# Spatial variation and correlates of raised blood pressure in Cambodia: a Gaussian-process geostatistical analysis of the 2023 STEPwise survey

**DOI:** 10.64898/2026.10.13.26365387

**Authors:** Samnang Um, Chhorvann Chhea, Maly Phy, Rei Haruyama

## Abstract

**Introduction:** Raised blood pressure is an important cardiovascular risk factor in Cambodia; however, national averages may obscure substantial local geographic heterogeneity. We assessed the spatial distribution of raised blood pressure and its demographic, metabolic, behavioural, built-environment, and environmental correlates using the 2023 Cambodia STEPwise approach to noncommunicable disease (NCD) risk factor surveillance (STEPS) survey.

**Methods:** We analysed 289 georeferenced primary sampling units (PSUs) from the 2023 Cambodia STEPS Survey. PSU-level case counts and numbers of adults with valid blood pressure data were modelled using a frequentist binomial generalised additive model with a two-dimensional Gaussian-process smooth. An explanatory model included selected PSU-level and contextual covariates; a separate mapping model used spatially continuous predictors to generate predictions on a 5-km grid. Design-based national and residence-specific prevalence estimates were calculated separately using the survey design and Step-2 weights.

**Results:** Among 4,225 adults with valid blood pressure data, unweighted prevalence of raised blood pressure was 28.6%, compared with 18.8% after Step-2 weighting; weighted prevalence was 17.5% in rural areas and 21.7% in urban/suburban areas. Mean observed unweighted PSU-level prevalence was 29.0%. In the adjusted spatial explanatory model, the proportion aged 60–69 years (β=1.969, SE=0.327; p<0.001) and overweight/obesity prevalence (β=0.996, SE=0.265; p<0.001) were positive ecological correlates. The spatial model had lower Akaike information criterion and residual deviance than its non-spatial counterpart, although Bayesian information criterion was higher. Prediction uncertainty was greater in some sparsely sampled areas.

**Conclusions:** Raised blood pressure showed significant geographic heterogeneity across Cambodia. Older age composition and overweight/obesity prevalence were positive PSU-level correlates, not individual-level causal effects. Predicted prevalence should be interpreted alongside uncertainty and should complement, rather than replace, design-based STEPS prevalence estimates for geographically informed NCD planning.

**WHAT IS ALREADY KNOWN ON THIS TOPIC:** Cambodia’s national STEPS Survey indicates a rising burden of raised blood pressure, but national averages cannot show fine-scale geographic heterogeneity or identify areas where model-based spatial estimates are least precise.

**WHAT THIS STUDY ADDS:** Using 289 georeferenced PSUs, this study distinguishes survey-weighted prevalence estimation from unweighted PSU-level geostatistical modelling, identifies older age composition and overweight/obesity as the strongest adjusted ecological correlates, and maps predicted prevalence alongside prediction standard errors and 95% confidence limits.

**HOW THIS STUDY MIGHT AFFECT RESEARCH, PRACTICE OR POLICY:** The maps can complement STEPS and routine NCD indicators by identifying geographic variations in the modelled raised blood pressure burden while explicitly identifying where the uncertainty is high. Areas with elevated predicted raised blood pressure prevalence and acceptable precision may warrant closer assessment of screening and treatment capacity, whereas areas with high uncertainty may benefit from additional surveillance.

## INTRODUCTION

Raised blood pressure is a major modifiable risk factor for cardiovascular disease, stroke, and premature mortality worldwide ^1-4^. Low- and middle-income countries bear a large share of this burden and continue to face substantial challenges in detection, treatment, and control ^1 4 5^. In Southeast Asia, population ageing, urbanisation, and the rising prevalence of overweight and obesity are likely to further increase the need for effective prevention, early detection, and long-term management of raised blood pressure.

Cambodia has experienced a substantial increase in the prevalence of raised blood pressure. The 2023 STEPwise Approach to the NCD Risk Factor Surveillance (STEPS) country report estimated a prevalence of 19.9% among adults aged 25–64 years, compared with 14.5% in 2016 and 11.2% in 2010 ^6^. The Ministry of Health has prioritised hypertension and diabetes screening and management in primary care through national standard operating procedures and implementation of the WHO Package of Essential Noncommunicable Disease Interventions (WHO PEN) ^7 8^.

National surveys such as STEPS provide representative estimates of non-communicable disease (NCD) risk factors and conditions; however, national or broad administrative summaries may obscure local geographic heterogeneity [6]. Geospatial analysis can complement design-based survey estimates by characterising spatial variation in the observed disease burden and relevant contextual exposures. Environmental and built-environment factors, including ambient PM2.5, urbanicity, geographic access to health services, and climatic conditions may vary spatially and contribute to cardiovascular risk or access to prevention and care ^9-12^. Geostatistical methods provide a framework for modelling such geographic variations while accounting for residual spatial dependence ^13-16^.

Despite the increasing use of geospatial methods in public health, fine-scale analyses of raised blood pressure remain limited in Cambodia. Therefore, this study aimed to (1) describe the geographic distribution of observed primary sampling unit (PSU)-level raised blood pressure; (2) assess demographic, metabolic, behavioural, built-environment, and environmental correlates using a binomial Gaussian-process spatial generalized additive model (GAM); and (3) generate a national 10-km prediction surface only using predictors with wall-to-wall spatial coverage. The analysis was designed to complement, rather than replace, survey-weighted national prevalence estimates ^13 14^.

## METHODS

### Study Setting

Cambodia is a lower-middle-income country in Southeast Asia, located between 10°N and 15°N latitude and 102°E and 108°E longitude, sharing borders with Thailand, Lao PDR and Vietnam, with a coastline along the Gulf of Thailand ^17^. The country has a tropical monsoon climate characterised by distinct wet and dry seasons that influence population health risks and environmental exposures relevant to cardiovascular disease. Cambodia is undergoing rapid demographic, economic, and epidemiological transitions characterised by increasing urbanisation, population aging, and a growing burden of NCDs ^8 17^.

### Data Source and Study Design

This study was conducted as a secondary geospatial analysis of the Cambodia STEPS Survey 2023 ^6^. The survey was implemented by the Department of Preventive Medicine, Ministry of Health, in coordination with the National Institute of Public Health, and with technical and financial support from the World Health Organization. The STEPS uses a multistage cluster sampling design to obtain nationally representative estimates of NCD risk factors. For the present spatial analysis, georeferenced PSUs were linked to participant-derived cluster-level summaries and environmental raster covariates. The final spatial dataset comprised 289 PSUs. Residence was classified for 288 PSUs (198 rural and 90 urban/suburban). One PSU lacked a residence classification and was retained in the analyses that did not require residence stratification.

### Outcome Measurement

The primary outcome was raised blood pressure among adults aged 18–69 years. Blood pressure was measured using the standardised STEPS protocol, with analytical blood pressure calculated from the second and third valid readings. Raised blood pressure was defined as systolic blood pressure ≥140 mmHg, diastolic blood pressure ≥90 mmHg, or current use of antihypertensive medication ^6^. For geostatistical modelling, individual-level outcomes were aggregated within each PSU to obtain the number of cases with raised blood pressure cases and the total number with valid blood pressure measurements ^6 16 18^.

### Geospatial Covariate Sourcing and Preprocessing

A multi-domain covariate database was assembled from participant-derived PSU summaries and spatially continuous environmental and built-environment data. The candidate domains included demographic composition, education, employment, overweight/obesity, smoking, alcohol consumption, nighttime lights, elevation, PM_2.5_, travel time to the nearest health facility, population density, heat-wave indicators, rainfall/precipitation, annual temperature, and urbanicity. Continuous raster layers were harmonised to a common coordinate reference system and linked to survey locations using 2-km buffers for urban clusters and 5-km buffers for rural clusters. Continuous exposures were summarised by the mean within each buffer, and categorical urbanicity was summarised using the modal class. Descriptive raster surfaces were retained to document the spatial coverage and geographic gradients of candidate contextual predictors (**Supplementary Figure 1**). Display transformations for highly skewed variables, including population density, travel time, nighttime lights, and target adult population, were applied solely for visualisation and did not alter the analytical covariates.

### Statistical and Geostatistical Analysis

#### 1. Survey-weighted prevalence and PSU-level analysis

Individual-level STEPS data were analysed using the survey design and Step-2 sampling weights to estimate population prevalence of raised blood pressure. For the descriptive spatial analyses and geostatistical models, observations were aggregated into unweighted raised blood pressure case counts and numbers of participants with valid outcome data within each PSU. Population-level weighted estimates were interpreted separately from the PSU-level model results. Detailed procedures for predictor screening, model fitting, spatial prediction and residual diagnostics follow.

#### 2. Multicollinearity Screening and Feature Selection

Before multivariable modelling, the candidate predictor set was examined using Pearson correlation and variance-inflation diagnostics. The full correlation matrix was examined to identify redundant representations of the same construct, while univariable scatterplots with locally weighted smoothing were used to assess the direction, approximate strength, and potential nonlinearity of crude PSU-level associations with observed raised blood pressure prevalence (**Supplementary Figures 2 and 3**). Variance inflation factor/generalized variance inflation factor (VIF/GVIF) was used as a diagnostic tool rather than for automatic variable selection. Highly redundant variables, particularly overlapping measures of temperature and age composition measures, were excluded from the adjusted model, whereas conceptually distinct predictors with moderate residual collinearity were retained and interpreted cautiously.

For transparency, the principal screening statistics are presented as:

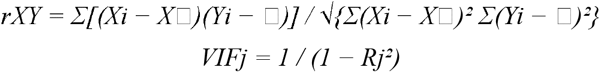

where *rXY* is the Pearson correlation coefficient, and *Rj*^*2*^ is the coefficient of determination obtained by regressing predictor *j* on the remaining predictors. VIF was used as a diagnostic indicator of multicollinearity rather than as evidence of a spatial effect.

#### 3. Geostatistical Modelling Framework

Cluster-level raised-blood-pressure counts were analysed using a frequentist binomial GAM fitted with the mgcv package (https://cran.r-project.org/web/packages/mgcv/index.html). For PSU *i*, the number of patients with raised blood pressure was modelled relative to the number of patients with valid blood pressure measurements. Fixed effects represented the selected cluster-level covariates, while residual spatial dependence was modelled using two-dimensional Gaussian-process smooths of longitude and latitude. Continuous predictors were standardised before estimation so that the fixed-effect coefficients represented changes in log odds per standard deviation increase in the corresponding covariate. This approach did not involve a Bayesian model ^5 11 13-16^.

The fitted cluster-level response is expressed as:

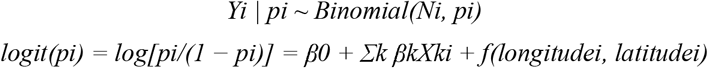

where *Yi* is the number of raised-blood-pressure cases in cluster i, *Ni* is the number of participants with valid blood-pressure measurements in that cluster, *pi* is the cluster-specific probability of raised blood pressure, β0 is the intercept, *βk* is the coefficient for covariate *Xki*, and *f(·)* is the two-dimensional Gaussian-process smooth fitted using mgcv.

The basis dimension *k*=60 controls the maximum complexity available for smoothing and is not interpreted as a spatial range parameter ^15 16^.

### National Risk Surface Interpolation and Uncertainty Tracking

Two related models were used for different purposes. The explanatory model included selected participant-derived cluster summaries together with environmental and built-environment covariates to estimate ecological associations at the PSU level. A separate national mapping model was fitted using predictors with continuous, wall-to-wall raster coverage across Cambodia. Participant-derived characteristics such as age composition, education, overweight/obesity, smoking, and alcohol use, were therefore not interpolated to unsampled locations. Predictions were generated on a 5-km analytical grid and back-transformed from the logit scale. The national prediction figure presents the mean predicted prevalence, prediction standard error on the response scale in percentage points, and the lower and upper 95% confidence limits. Any finer resampling used for figure rendering was purely visual and did not alter the analytical grid or numerical inference.

The predictions on the logit scale were transformed into prevalence probabilities using the following inverse logit function:

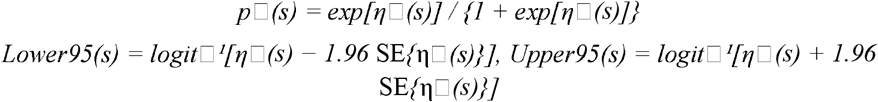

where η□*(s)* is the predicted linear predictor at location *s*, and SE{η□*(s)}* is its model-based standard error on the link scale. Lower and upper confidence limits are calculated on the link scale and transformed using the inverse-logit function. For the geographic presentation of the uncertainty, the prediction standard error is also expressed on the response probability scale and reported as a percentage point.

### Residual Autocorrelation and Validation Diagnostics

Model fit was evaluated using residual deviance, deviance explained, Akaike information criterion (AIC), and Bayesian information criterion (BIC). The non-spatial and spatial explanatory models used the same fixed-effect structure, allowing assessment of the contribution of the Gaussian-process smooth while recognising that different information criteria may favour different levels of model complexity. Residual spatial structure was assessed using an empirical variogram of Pearson residuals with a 95% Monte Carlo permutation envelope. The variogram was interpreted as a diagnostic evaluation of the remaining spatial dependence rather than as evidence of complete spatial independence ^5 11 13 14 16^.

For residual spatial diagnostics, the empirical semivariogram at lag *h* is calculated as:

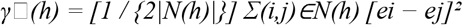

where *N(h)* is the set of cluster pairs separated by distances within lag class *h*; |*N(h)*| is the number of such pairs; and *ei* and *ej* are the model residuals at locations *i* and *j*, respectively. The Monte Carlo envelope is obtained by repeatedly permuting the residuals across locations and calculating the 2.5^th^ and 97.5^th^ percentiles of semivariance within each lag class.

### Ethical Considerations

The Cambodia STEPS Survey 2023 was approved by the National Ethics Committee for Health Research (NECHR), Ministry of Health, Cambodia (No. 116 NECHR, 11 April 2023). As this study used anonymised secondary data, additional ethical approval was not required.

## RESULTS

### Descriptive Characteristics of the Spatial Clusters

In total, 289 georeferenced PSUs were included in the spatial analysis. Residence data were available for 288 PSUs, comprising 198 rural and 90 urban/suburban PSUs; one PSU had an unclassified residence. The cleaned individual-level dataset included 4,225 adults with valid blood pressure measurements. **Figure 1** shows the geographic distribution of the sampled PSUs and the observed, unweighted PSU-level prevalence of raised blood pressure. The model-based prevalence and uncertainty surfaces are presented in **Figure 2**, and the residual spatial diagnostic is presented in **Figure 3**; their results are discussed in the corresponding subsections below.

**Figure 1.**
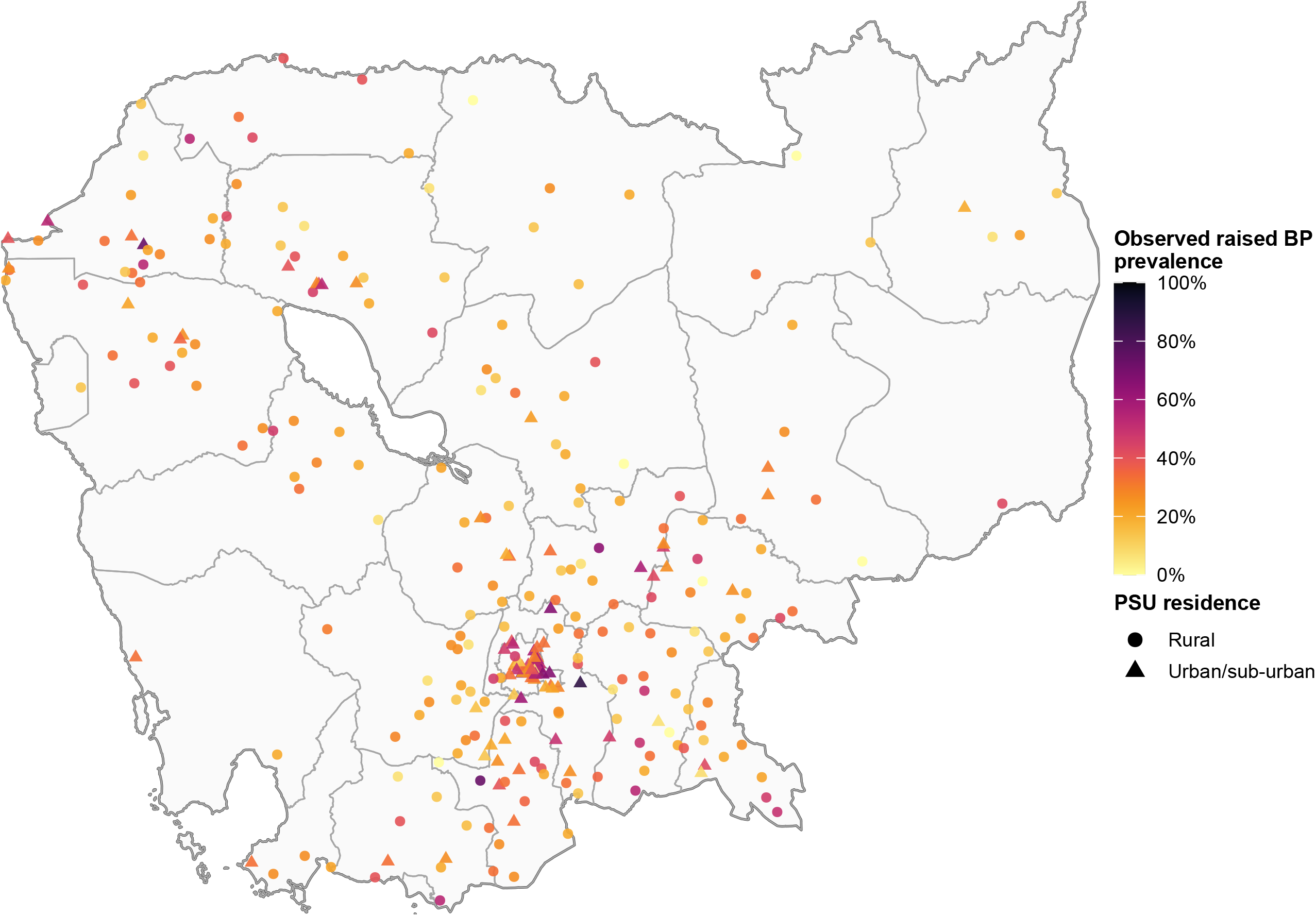
Geographic distribution of sampled primary sampling units (PSUs) and observed raised blood pressure prevalence, Cambodia STEPS Survey 2023. Point colour represents the unweighted observed PSU-level prevalence of raised blood pressure (0– 100%), and point shape distinguishes rural and urban/suburban PSUs where residence was classified. Province boundaries are shown for geographic reference.

**Figure 2.**
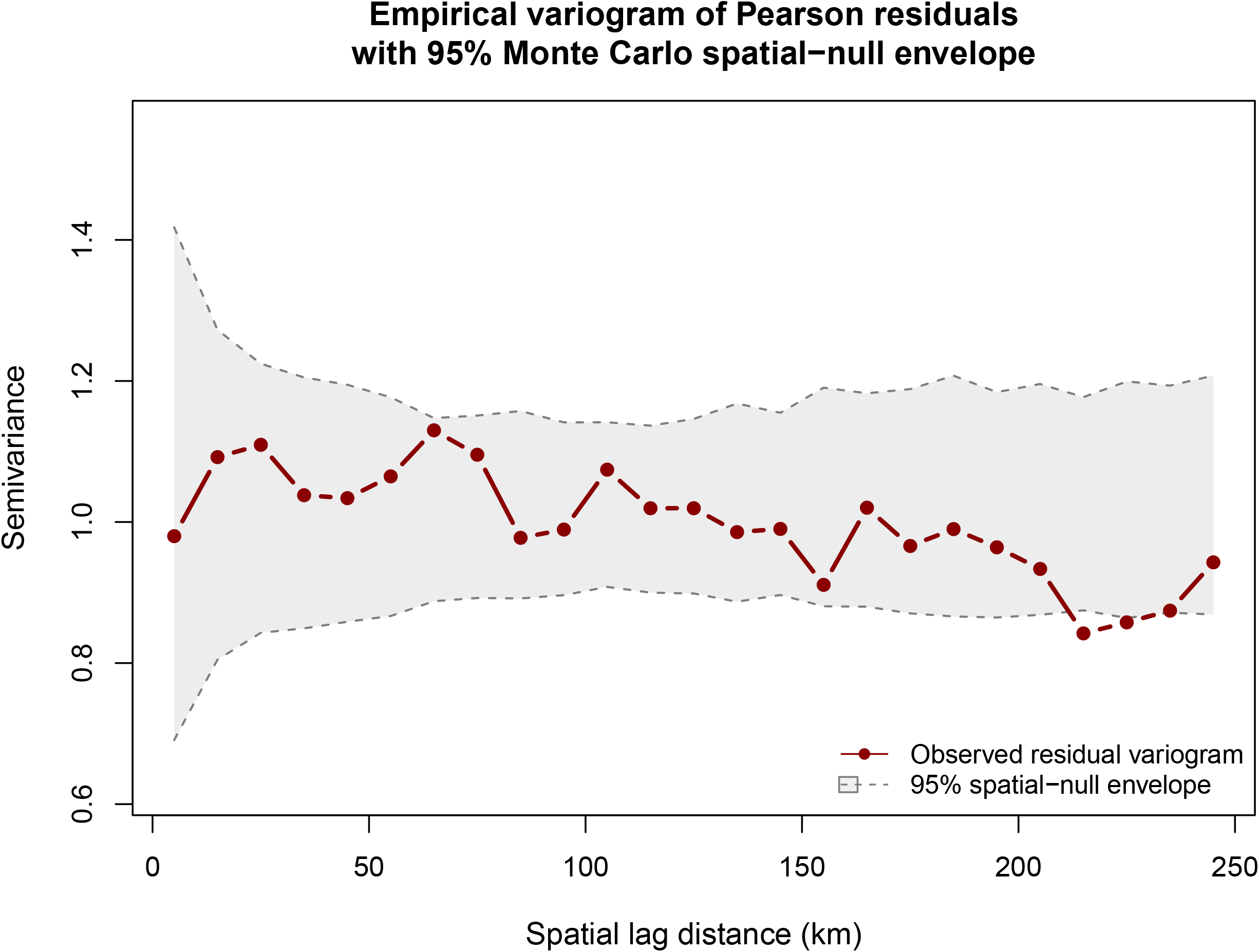
Empirical variogram of Pearson residuals with a 95% Monte Carlo spatial-null envelope. The observed residual semivariance is plotted across spatial lag distances and compared with the 2.5^th^ and 97.5^th^ percentile envelope obtained from 199 random permutations of residuals across primary sampling unit (PSU) locations. The observed variogram was largely contained within the envelope, failing to provide strong evidence of substantial residual spatial autocorrelation after adjustment.

**Figure 3.**
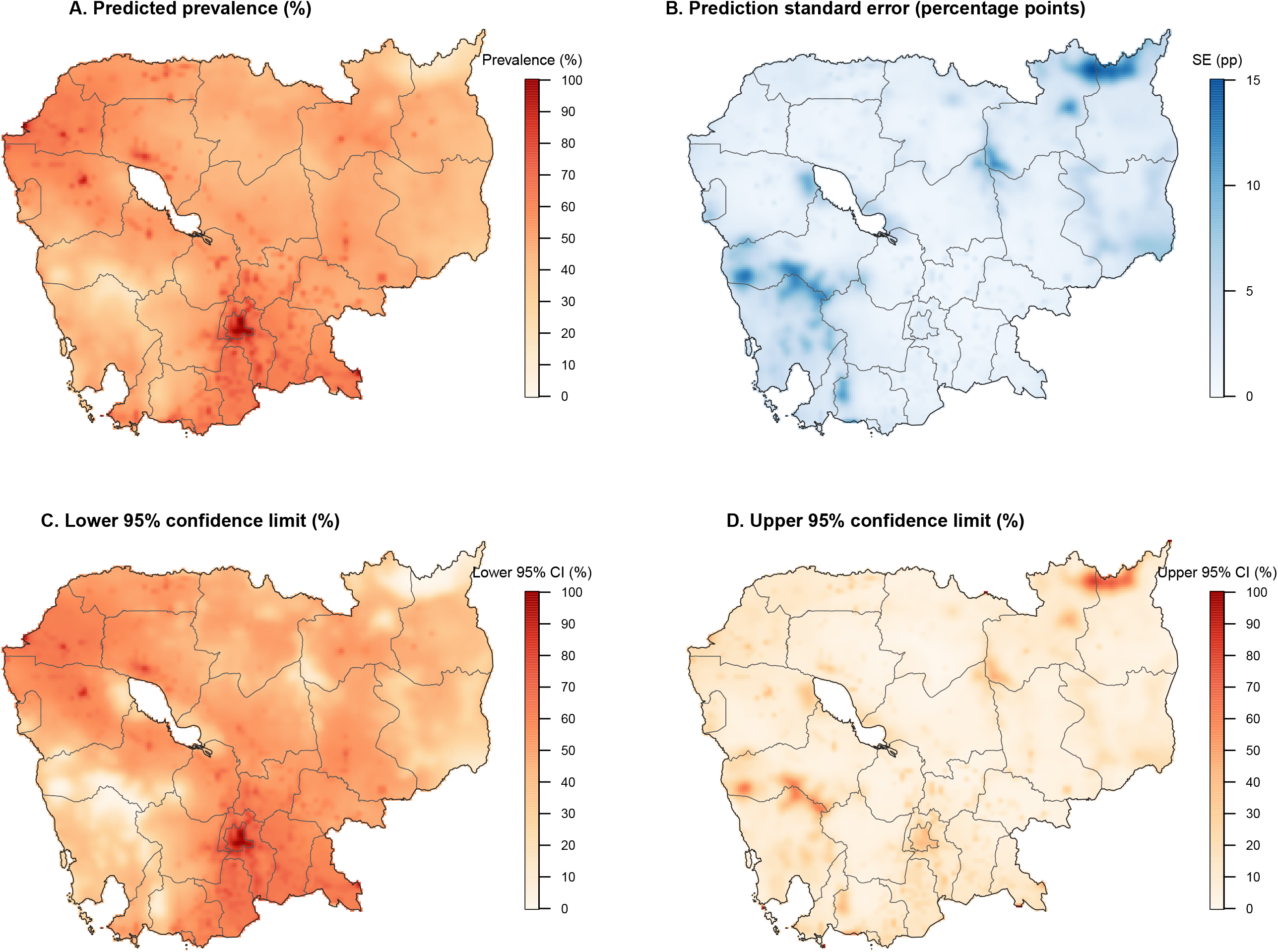
Spatial distribution of model-predicted raised blood pressure prevalence and associated uncertainty among adults aged 18–69 years in Cambodia, using the 2023 STEPS survey. (**A**) Predicted prevalence (%); (**B**) prediction standard error (percentage points); (**C**) lower 95% confidence limit (%); and (**D**) upper 95% confidence limit (%). Estimates were generated using a binomial spatial generalised additive model on a 5-km analytical grid. Provincial boundaries are shown for geographic reference.

The distinction between weighted population prevalence and unweighted spatial-model outcomes was substantial. Among adults with valid blood pressure data, the unweighted prevalence was 29.0%, whereas the Step-2 weighted prevalence was 18.8%. The weighted prevalence was 17.5% among rural participants and 21.7% among urban/suburban participants. These weighted estimates are appropriate for describing population-level prevalence. The higher unweighted cluster/participant proportions were used solely as inputs to the binomial geostatistical analysis and should not be interpreted as nationally representative prevalence estimates ^6 19^.

**Table 1** summarises the unweighted PSU-level characteristics and descriptive rural– urban comparisons. The mean observed PSU-level prevalence of raised blood pressure was 29.0% overall, 25.6% in rural PSUs, and 36.5% in urban/suburban PSUs (p<0.001). The mean age was 46.5 years overall and was higher in urban/suburban than in rural PSUs (47.9 versus 45.9 years; p=0.002). The mean years of schooling was also higher in urban/suburban PSUs (6.3 versus 4.7 years; p<0.001), whereas no formal schooling was more common in rural PSUs (18.5% versus 11.8%; p<0.001). These p-values represent descriptive PSU-level comparisons and should not be interpreted as substitutes for survey-design-based population inference.

**Table 1.** Descriptive characteristics of sampled PSUs, observed raised blood pressure prevalence, and rural–urban comparisons.

| Characteristics / Covariate Dimension | Overall Mean / Proportion (n=289) | Rural Clusters (n=198) | Urban Clusters(n=90) | p-value |
| --- | --- | --- | --- | --- |
| Observed raised blood pressure prevalence among sampled PSUs (%) | 29.0% | 25.6% | 36.5% | <0.001 |
| Proportion of Female (%) | 64.4% | 63.8% | 65.8% | 0.246 |
| Mean Age (Years) | 46.5 | 45.9 | 47.9 | 0.002 |
| Age 18–29 Years (%) | 13.3% | 14.2% | 11.6% | 0.048 |
| Age 30–39 Years (%) | 19.1% | 20.2% | 16.8% | 0.022 |
| Age 40–49 Years (%) | 21.4% | 21.7% | 20.8% | 0.557 |
| Age 50–59 Years (%) | 24.2% | 23.5% | 25.6% | 0.165 |
| Age 60–69 Years (%) | 22.0% | 20.4% | 25.1% | 0.006 |
| Mean Schooling (Years) | 5.2 | 4.7 | 6.3 | <0.001 |
| No Formal Schooling (%) | 16.4% | 18.5% | 11.8% | <0.001 |
| Completed Primary (%) | 48.4% | 51.0% | 42.3% | <0.001 |
| Completed at least Secondary (%) | 35.3% | 30.5% | 45.9% | <0.001 |
| Currently in Marital (%) | 68.9% | 71.3% | 63.4% | <0.001 |
| Previously Married (%) | 22.3% | 20.8% | 25.4% | 0.011 |
| Never Married (%) | 8.9% | 7.8% | 11.1% | 0.002 |
| Gainfully Employed (%) | 77.0% | 78.2% | 74.5% | 0.050 |
| Working from Home (%) | 16.5% | 16.0% | 17.4% | 0.397 |
| Actively Unemployed (%) | 4.5% | 4.3% | 4.9% | 0.441 |
| Outside Active Labor Force (%) | 2.1% | 1.6% | 3.4% | 0.003 |
| Overweight and Obesity (%) | 30.6% | 27.1% | 38.0% | <0.001 |
| Current Tobacco Smoking (%) | 15.8% | 17.3% | 12.5% | <0.001 |
| Current Alcohol Consumption (%) | 73.5% | 74.1% | 72.1% | 0.353 |
| Nighttime Light Intensity (0–1) | 0.3 | 0.1 | 0.7 | <0.001 |
| Elevation Above Sea Level (m) | 39.7 | 47.3 | 23.5 | <0.001 |
| Fine Particulate Matter Exposure (PM<sub>2.5</sub>, µg/m<sup>3</sup>) | 19.6 | 19.2 | 20.3 | <0.001 |
| Travel Time to Nearest Health Facility (minutes) | 30.9 | 39.6 | 12.0 | <0.001 |
| Population Density (people/km<sup>2</sup>) | 2134.8 | 295.8 | 6188.7 | <0.001 |
| Extreme Heatwave Index Score | 27.4 | 28.8 | 24.3 | 0.015 |
| Annual Heatwave Frequency (Events/Year) | 8.3 | 8.3 | 8.2 | 0.494 |
| Average Annual Rainfall (mm/year) | 1577.6 | 1618.6 | 1490.9 | 0.008 |
| Annual Precipitation 2020 (mm) | 1621.9 | 1657.5 | 1547.3 | 0.035 |
| Annual Maximum Temperature (°C) | 33.3 | 33.2 | 33.4 | 0.080 |
| Annual Minimum Temperature (°C) | 25.2 | 25.1 | 25.5 | <0.001 |
| Average Annual Temperature (°C) | 29.2 | 29.2 | 29.5 | <0.001 |

**Note:** Values are unweighted PSU-level means/proportions. p-values compare rural versus urban/sub-urban PSUs using Welch's two-sample t-test and are descriptive only. The raised blood pressure row is the mean of PSU-specific observed prevalences, not the survey-weighted national prevalence. One of 289 PSUs had unclassified residence and therefore did not contribute to the rural–urban comparison. National and residence-specific population prevalence should be reported from individual-level STEPS analysis using the survey design and Step-2 weights. p-values are reported to three decimals; values <0.001 are shown as <0.001.

Metabolic and behavioural characteristics also differed between residential groups. The mean PSU-level prevalence of overweight/obesity was higher in urban/suburban PSUs than in rural PSUs (38.0% vs. 27.1%, p<0.001), whereas current tobacco smoking was more prevalent in rural PSUs (17.3% vs. 12.5%, p<0.001). Current alcohol consumption was high in both groups and did not differ significantly at the PSU level (74.1% vs. 72.1%; p=0.353).

Several contextual exposures differed markedly by residence. The mean travel time to the nearest health facility was 39.6 min in rural PSUs and 12.0 min in urban/suburban PSUs (p<0.001). The average population density was 295.8/km^2^ in rural PSUs and 6,188.7/km^2^ in urban/suburban PSUs (p<0.001). Mean PM_2.5_ exposure was 19.2 versus 20.3 µg/m3 (p<0.001), whereas annual rainfall was higher in rural PSUs (1,618.6 vs. 1,490.9 mm/year; p=0.008).

### Correlation, Univariable Patterns, and Multicollinearity

The correlation heatmap showed that the prevalence of PSU-level raised blood pressure had the strongest positive crude correlations with mean age (r=0.46), proportion of patients aged 60–69 years (r=0.42), overweight/obesity prevalence (r=0.39), and nighttime-light intensity (r=0.38). Negative crude correlations were observed with the proportion aged 18–29 years (r=-0.35), current tobacco smoking (r=-0.28), and travel time to the nearest health facility (r=-0.25). Population density showed a modest positive correlation (r=0.26), whereas several climatic indicators were only weakly correlated with observed prevalence (**Supplementary Figure 3**).

The univariable scatterplots were consistent with these findings and suggested nonlinear relationships for several predictors (**Supplementary Figure 2**). The mean age and the proportion of patients aged 60–69 years showed progressively higher observed prevalence toward the upper end of their distributions, whereas overweight/obesity prevalence showed a broadly positive association. Nighttime-light intensity and population density also showed positive crude gradients, whereas travel time and current tobacco smoking showed inverse crude trends. These plots are descriptive and did not account for confounding factors, ecological aggregation or residual spatial dependence; therefore, they were used to support the diagnostics and interpretation rather than as stand-alone evidence of independent associations.

The predictor–predictor correlation was substantial for several overlapping constructs. The strongest redundancy occurred among the annual temperature indicators and the mutually constrained demographic composition measures. These patterns were consistent with the VIF/GVIF results and supported the exclusion of redundant temperature and age representations prior to fitting of the adjusted model. Moderate residual collinearity among the selected, conceptually distinct predictors was retained and treated as a limitation, necessitating cautious interpretation of the coefficients.

### Spatial Distribution of Contextual Predictors

The descriptive spatial predictor matrix demonstrated substantial geographic variation in the contextual inputs used during model development (**Supplementary Figure 1**). Elevation was greatest in western and northeastern upland areas, rainfall and precipitation showed broad regional gradients, and travel time to the nearest health facility was generally longer in peripheral and less densely populated areas than around the central urban corridor. Population density and nighttime-light intensity were strongly concentrated around Phnom Penh and other urbanised locations, whereas much of the country had low population density and low nighttime-light values. These maps illustrate the spatial coverage and heterogeneity of the candidate covariates rather than depicting the independent effects of raised blood pressure.

### Model Performance and Spatial Validation

The goodness-of-fit diagnostics are summarised in **Supplementary Table 2**. Compared with the non-spatial explanatory generalized linear model, the spatial GAM reduced the AIC from 1177.67 to 1151.45, and the residual deviance from 362.20 to 302.89, while deviance explained increased from 15.8% to 37.7%. In contrast, the BIC increased from 1203.29 to 1237.60, reflecting the greater complexity of the spatial model. Taken together, these diagnostic findings indicate improved descriptive fit according to AIC and deviance-based criteria but do not support the description of the spatial model as uniformly superior across all information criteria.

The empirical variogram of the Pearson residuals was largely contained within the 95% Monte Carlo spatial null envelope over lag distances of approximately 0–250 km (**Figure 2**). Observed semivariance fluctuated around approximately 0.9–1.1 across most distance classes without a sustained increase beyond the permutation envelope. This pattern did not provide strong evidence of substantial residual spatial autocorrelation after adjustment. The variogram was used as a diagnostic tool rather than as evidence of complete spatial independence, and minor departures in individual lag classes were not overinterpreted.

### Adjusted Geostatistical Estimates and Vulnerability Drivers

The fixed effects estimates from the crude and adjusted geostatistical models are presented in **Table 2**. In the adjusted explanatory model, the strongest positive cluster-level associations were observed for the proportion of adults aged 60–69 years and prevalence of overweight/obesity. Because these covariates were aggregated at the PSU level, the estimates represent ecological associations and not individual-level causal effects.

**Table 2.**
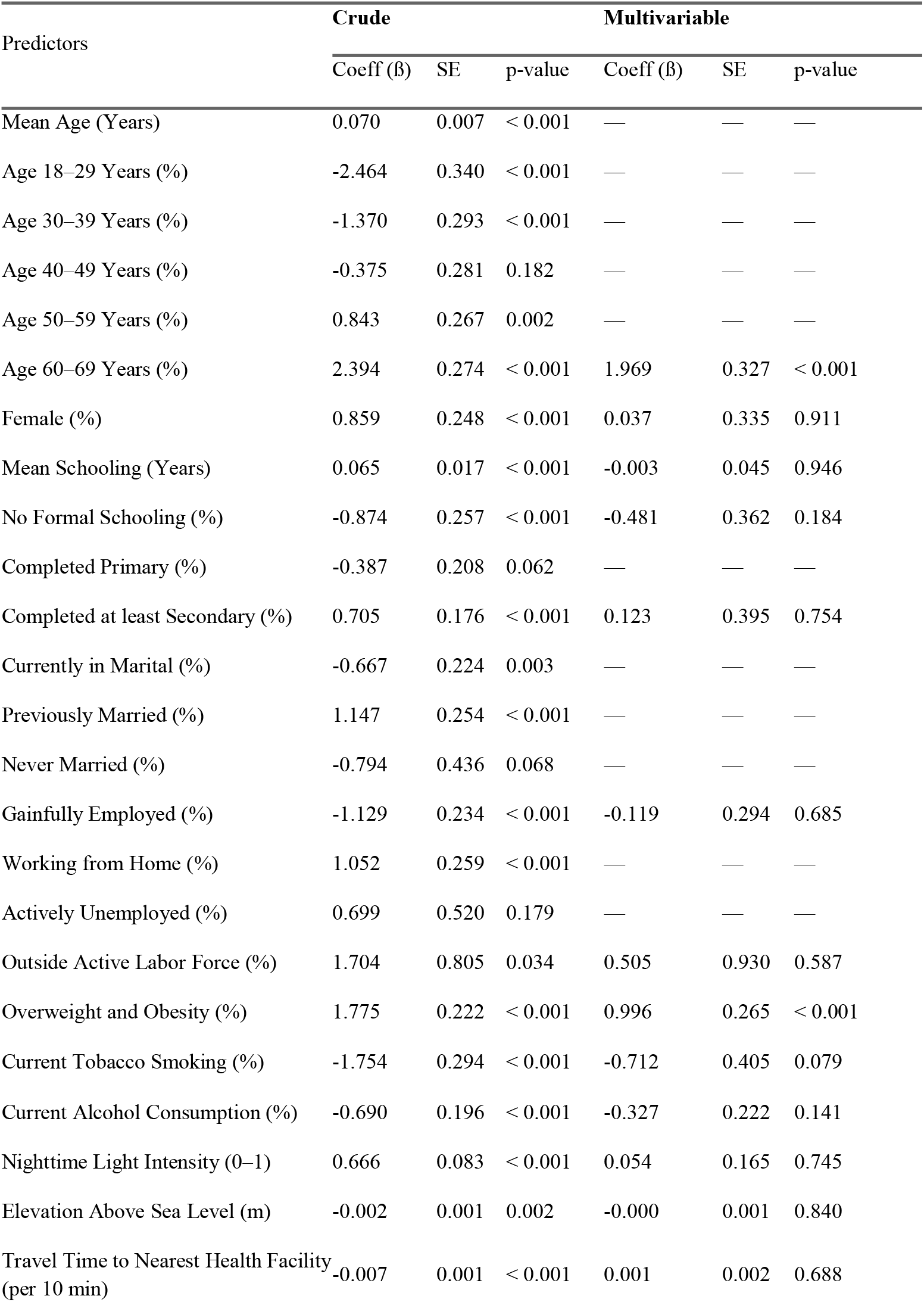
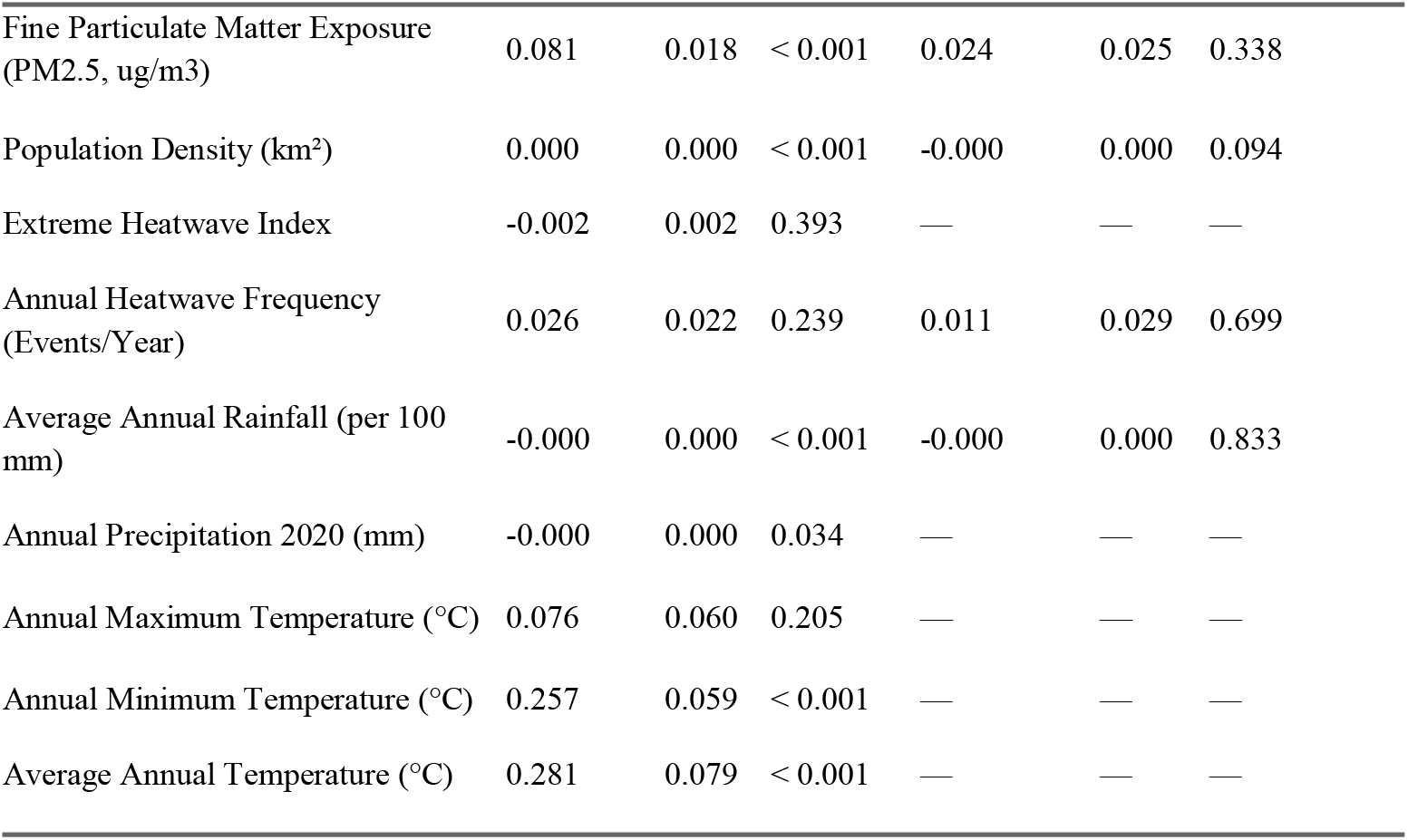
Crude and Multivariable Geostatistical Parameter Estimates Matrix.

| Predictors | Crude |  |  | Multivariable |  |  |
| --- | --- | --- | --- | --- | --- | --- |
|  | Coeff (ß) | SE | p-value | Coeff (ß) | SE | p-value |
| Mean Age (Years) | 0.070 | 0.007 | < 0.001 | — | — | — |
| Age 18–29 Years (%) | -2.464 | 0.340 | < 0.001 | — | — | — |
| Age 30–39 Years (%) | -1.370 | 0.293 | < 0.001 | — | — | — |
| Age 40–49 Years (%) | -0.375 | 0.281 | 0.182 | — | — | — |
| Age 50–59 Years (%) | 0.843 | 0.267 | 0.002 | — | — | — |
| Age 60–69 Years (%) | 2.394 | 0.274 | < 0.001 | 1.969 | 0.327 | < 0.001 |
| Female (%) | 0.859 | 0.248 | < 0.001 | 0.037 | 0.335 | 0.911 |
| Mean Schooling (Years) | 0.065 | 0.017 | < 0.001 | -0.003 | 0.045 | 0.946 |
| No Formal Schooling (%) | -0.874 | 0.257 | < 0.001 | -0.481 | 0.362 | 0.184 |
| Completed Primary (%) | -0.387 | 0.208 | 0.062 | — | — | — |
| Completed at least Secondary (%) | 0.705 | 0.176 | < 0.001 | 0.123 | 0.395 | 0.754 |
| Currently in Marital (%) | -0.667 | 0.224 | 0.003 | — | — | — |
| Previously Married (%) | 1.147 | 0.254 | < 0.001 | — | — | — |
| Never Married (%) | -0.794 | 0.436 | 0.068 | — | — | — |
| Gainfully Employed (%) | -1.129 | 0.234 | < 0.001 | -0.119 | 0.294 | 0.685 |
| Working from Home (%) | 1.052 | 0.259 | < 0.001 | — | — | — |
| Actively Unemployed (%) | 0.699 | 0.520 | 0.179 | — | — | — |
| Outside Active Labor Force (%) | 1.704 | 0.805 | 0.034 | 0.505 | 0.930 | 0.587 |
| Overweight and Obesity (%) | 1.775 | 0.222 | < 0.001 | 0.996 | 0.265 | < 0.001 |
| Current Tobacco Smoking (%) | -1.754 | 0.294 | < 0.001 | -0.712 | 0.405 | 0.079 |
| Current Alcohol Consumption (%) | -0.690 | 0.196 | < 0.001 | -0.327 | 0.222 | 0.141 |
| Nighttime Light Intensity (0–1) | 0.666 | 0.083 | < 0.001 | 0.054 | 0.165 | 0.745 |
| Elevation Above Sea Level (m) | -0.002 | 0.001 | 0.002 | -0.000 | 0.001 | 0.840 |
| Travel Time to Nearest Health Facility (per 10 min) | -0.007 | 0.001 | < 0.001 | 0.001 | 0.002 | 0.688 |
|  | Coeff (β) | SE | p-value | Coeff (β) | SE | p-value |
| Fine Particulate Matter Exposure (PM2.5, ug/m3) | 0.081 | 0.018 | < 0.001 | 0.024 | 0.025 | 0.338 |
| Population Density (km<sup>2</sup>) | 0.000 | 0.000 | < 0.001 | -0.000 | 0.000 | 0.094 |
| Extreme Heatwave Index | -0.002 | 0.002 | 0.393 | — | — | — |
| Annual Heatwave Frequency (Events/Year) | 0.026 | 0.022 | 0.239 | 0.011 | 0.029 | 0.699 |
| Average Annual Rainfall (per 100 mm) | -0.000 | 0.000 | < 0.001 | -0.000 | 0.000 | 0.833 |
| Annual Precipitation 2020 (mm) | -0.000 | 0.000 | 0.034 | — | — | — |
| Annual Maximum Temperature (°C) | 0.076 | 0.060 | 0.205 | — | — | — |
| Annual Minimum Temperature (°C) | 0.257 | 0.059 | < 0.001 | — | — | — |
| Average Annual Temperature (°C) | 0.281 | 0.079 | < 0.001 | — | — | — |

A higher cluster proportion of adults aged 60–69 years was positively associated with hypertension (β=1.969, SE=0.327, p<0.001). As continuous predictors were standardised in the R model, this coefficient corresponded to a 1-SD increase in the cluster-level age proportion rather than a one-percentage point increase. A higher cluster-level prevalence of overweight/obesity was also positively associated with raised blood pressure (β=0.996, SE=0.265, p<0.001). This ecological association is consistent with the positive crude relationship shown in **Supplementary Figure 2** but should not be interpreted as an individual-level effect.

Several behavioural variables associated with raised blood pressure in the crude models were attenuated after multivariable and spatial adjustment. Current smoking (β=-0.712, p=0.079) and current alcohol consumption (β=-0.327, p=0.141) were not statistically significant in the adjusted model. This attenuation may reflect confounding, collinearity, ecological aggregation, or spatial structure and should not be considered to indicate that these behaviours are unimportant individual-level cardiovascular risk factors. The adjusted associations for nighttime lights, elevation, travel time, PM_2.5_, population density, heatwave frequency, and rainfall were not statistically significant. However, these findings do not indicate the absence of environmental effects. Spatial scale, temporal alignment of raster products, exposure measurement error, residual collinearity, and limited number of PSUs may have reduced the power to detect independent associations after spatial smoothing [5,11,13,14].

### National Risk Surface Projections and Uncertainty Tracking

The national prediction and uncertainty surfaces from the raster-supported mapping model are shown in **Figure 3**. The predicted prevalence of raised blood pressure prevalence varied geographically, with most areas showing moderate estimated prevalence and several localised areas with higher predicted values. The prediction standard errors were spatially heterogeneous and were the greatest in several peripheral and sparsely sampled locations, particularly in parts of the northeast. The lower and upper 95% confidence limit surfaces were also wider in the same areas, indicating greater uncertainty and cautioning against interpreting apparent high- or low-prevalence areas as precise hotspot boundaries. The mapping model was intentionally restricted to predictors with wall-to-wall raster support and therefore differed from the explanatory model used to estimate PSU-level ecological associations.

To facilitate geographic interpretation at a finer administrative scale, the same four model-based surfaces are displayed with district administrative boundaries in **Supplementary Figure S4**. This presentation reveals within-district variation that may be obscured by province-only outlines, while retaining the same underlying 5-km predictions. Importantly, district lines are reference overlays and do not indicate separate district-level model fitting, district-specific observed prevalence, or formally aggregated district estimates. Apparent differences across district boundaries should therefore not be interpreted as discontinuities in risk.

The four panels should be read together rather than interpreting the predicted mean alone. Where the standard error is larger or the lower and upper confidence limits are widely separated, the data and fitted model support a broader range of plausible local prevalence values. Such locations may be priorities for improving geographic sampling or validating model predictions against independent data, but they cannot be designated high-burden areas with confidence solely on the basis of their mapped mean. The detailed district-boundary presentation in **Supplementary Figure S4** is intended to support visual assessment of geographic patterns, not to provide district-level estimates for ranking or resource allocation.

Prediction precision was generally heterogeneous and higher in areas with greater sampling support and lower in several peripheral sparsely sampled locations. Accordingly, the predicted prevalence surface should be interpreted jointly with Panel B and the upper and lower 95% confidence limit panels. Areas with a high predicted prevalence but large standard errors or wide confidence limits represent uncertain model-based estimates and may be prioritized for additional surveillance rather than definitive high-burden hotspots.

The raster-supported environmental and built-environment covariates used for national mapping are presented in **Supplementary Figure 1**. Participant-derived cluster variables were retained for explanatory modelling but were not interpolated across unsampled areas for the national prediction surface.

## DISCUSSION

Supplementary spatial diagnostics supported the interpretation of the main geostatistical findings. The predictor maps showed that healthcare accessibility, population density, nighttime lights, rainfall, elevation, and temperature were spatially structured rather than randomly distributed across Cambodia. Simultaneously, the correlation matrix showed substantial correlations among several contextual variables and demographic composition. Together, these findings reinforce the importance of distinguishing crude spatial patterns from independently adjusted associations.

The crude correlation pattern was dominated by age structure and metabolic composition. The mean age (r=0.46), proportion of patients aged 60–69 years (r=0.42), and overweight/obesity prevalence (r=0.39) showed the strongest positive correlations with the observed PSU-level prevalence. These findings were consistent with the adjusted model, in which the proportion of individuals aged 60–69 years and overweight/obesity prevalence remained the strongest positive ecological correlates. In contrast, crude associations with nighttime lights, population density, travel time, smoking, and several environmental variables were attenuated after multivariable and spatial adjustment, suggesting that part of their univariable associations may have reflected confounding, correlated contextual conditions, or shared spatial patterning.

### Principal Findings and Spatial Heterogeneity

This study demonstrates substantial geographic heterogeneity in raised blood pressure across Cambodia while also highlighting the importance of distinguishing design-based population prevalence from unweighted PSU observations. In the cleaned individual-level dataset, the Step-2 weighted prevalence was 18.8%, compared with an unweighted individual-level prevalence of 28.6% and a mean observed PSU prevalence of 29.0%. The weighted prevalence was 21.7% in urban/suburban areas and 17.5% in rural areas. The official STEPS report documents a substantial national burden and an increasing trend across successive survey rounds ^6 13 14 19^. The geostatistical analysis should therefore be interpreted as a complementary assessment of spatial heterogeneity and ecological correlates rather than as an alternative estimator of national prevalence.

The prediction maps provide information that cannot be obtained from a single national average by showing geographic variation in both modelled prevalence and prediction precision. However, the uncertainty surfaces are essential for interpretation. Several peripheral areas had comparatively large prediction standard errors and wide confidence limits, reflecting weaker local information. These areas should not be classified as definitive hotspots solely from the mean prediction surface; rather, they may identify areas where additional surveillance could improve spatial precision ^13 14 19^.

### Demographic Aging and Metabolic Transition

In the adjusted geostatistical model, clusters with a greater proportion of adults aged 60– 69 years had a higher prevalence of raised blood pressure prevalence. This finding is epidemiologically plausible given the established increase in blood pressure with age; however, the cluster-level association should not be interpreted as an individual-level causal effect. Rather, the results primarily identify areas in which an older local age structure coincided with a greater burden of raised blood pressure ^2 15 16 18 19^.

Cluster-level overweight/obesity prevalence showed the strongest positive adjusted correlation. Urban/suburban areas also showed higher overweight/obesity prevalence in the descriptive data. Together, these results are consistent with a metabolic transition in which areas with a higher concentration of excess body weight may experience a greater burden of raised blood pressure However, the cross-sectional ecological design precludes the establishment of temporality or individual-level mediation ^2 15 16 18 19^.

The combined pattern of an older local age structure and higher prevalence of overweight/obesity is consistent with the demographic and metabolic transitions accompanying NCD growth ^2 15 16 18 19^. The interpretation remains ecological: the coefficients indicate that PSUs with these characteristics tended to have a higher prevalence of raised blood pressure prevalence after adjustment, rather than demonstrating individual-level causal effects.

Smoking and alcohol consumption were not statistically significant after multivariable and spatial adjustment, despite stronger crude associations. This attenuation should be interpreted cautiously because behavioural exposures were summarised at the cluster level and may be affected by ecological measurement errors, residual confounding, and correlations with demographic or spatial characteristics ^2 18 20^.

Nighttime light, elevation, travel time to a health facility, PM2.5, population density, heat-wave frequency, and rainfall were not statistically significant in the adjusted explanatory model. These null cluster-level associations should also be interpreted cautiously because exposure measurement error, spatial scale, temporal mismatch among raster products, residual collinearity, and limited statistical power may have attenuated the associations ^9-12.^

### Alignment with the National Strategy and WHO PEN Framework

Cambodia has incorporated hypertension and diabetes screening and management into primary healthcare and has adopted approaches aligned with WHO PEN ^6 7^. Geospatial outputs can complement routine national and provincial indicators by identifying geographic variation in modelled burden and by revealing where predictions are less precise. For program planning, areas with elevated predicted prevalence and comparatively low prediction errors may warrant closer assessment of screening and follow-up capacity, whereas areas with high uncertainty may benefit from strengthened surveillance or additional data collection.

Spatial outputs can support WHO PEN planning by highlighting geographic variations in the modelled burden and areas of high prediction uncertainty. Policy use should prioritise locations where an elevated predicted prevalence is accompanied by reasonably narrow uncertainty intervals, whereas areas with wide intervals may warrant additional surveillance or targeted data collection before resource-allocation decisions are made ^6^.

### Strengths and Limitations

This study has several strengths. First, it uses nationally collected STEPS data, standardised blood pressure measurements, georeferenced PSUs, a binomial model that preserves the number of cases and valid examinations within each PSU, and explicit two-dimensional Gaussian-process spatial smoothing. It also distinguishes design-based prevalence estimation from spatial modelling and an explanatory model from a raster-supported national mapping model and presents the predicted prevalence together with response-scale prediction standard errors and 95% confidence limits ^6 13 14 16^.

This study also has some limitations. First, its cross-sectional design precludes causal inferences and temporal ordering. Second, the regression coefficients represent ecological PSU-level associations that cannot be directly transferred to individuals. Third, the correlation heatmap and VIF/GVIF diagnostics demonstrated residual collinearity among several candidate predictors, particularly overlapping temperature, demographic, and socioeconomic measures, which may reduce coefficient stability. Fourth, environmental rasters differed in reference period and spatial resolution and may imperfectly represent individual-level exposures. Fifth, the 10-km prediction grid provides model-based spatial estimates rather than direct observations, and the prediction standard error is greater when local sampling and covariate support are weaker. Sixth, the log- or square-root transformations used in the selected supplementary maps are display-only transformations and should not be interpreted as transformations of the fitted model. Finally, national and residence-specific population prevalence estimates must be obtained using the STEPS survey design and sampling weights, rather than the unweighted PSU data used for spatial mapping ^6 13 14 16^.

## CONCLUSIONS

Raised blood pressure in Cambodia shows substantial geographic heterogeneity; however, the population burden is best characterised using survey-weighted individual-level estimates. In the cleaned analytical dataset, the Step-2 weighted prevalence was 18.8%. At the PSU level, older age and overweight/obesity prevalence were the strongest adjusted positive ecological correlates. National prediction maps may support geographically informed NCD planning when the predicted prevalence is interpreted alongside prediction standard errors and 95% confidence limits.

For WHO PEN implementation and national NCD planning, geostatistical outputs should complement, rather than replace, existing surveys and administrative indicators. Locations with consistently elevated predicted prevalence and acceptable prediction precision may warrant an intensified assessment of screening, diagnosis, treatment, and follow-up capacity, whereas areas with high uncertainty may benefit from additional surveillance and data collection ^6-8^. Future research should validate the predicted geographic patterns using additional population-based data and examine whether areas with elevated raised blood pressure prevalence also experience gaps in screening, diagnosis, treatment and control.

## Data Availability

The dataset analyzed during the current study is based on the 2023 Cambodia STEPwise approach to noncommunicable disease (NCD) risk factor surveillance (STEPS) survey, implemented by the Ministry of Health of Cambodia and the World Health Organization. The de-identified dataset was submitted to WHO headquarters in September 2024 to be made publicly available on the WHO NCD Microdata Repository. Although the dataset is pending public appearance on the repository, it can be obtained from the Ministry of Health Cambodia upon reasonable request.

https://extranet.who.int/ncdsmicrodata/index.php/catalog/STEPS

## ACKNOWLEDGMENTS

The authors acknowledge the Department of Preventive Medicine, Ministry of Health of Cambodia, and the National Institute of Public Health for their contributions to the Cambodia STEPS 2023 survey. We thank the survey participants, field data collectors, supervisors, and technical teams whose efforts made this study possible. We also thank Editage for English-language editing support.

## CONTRIBUTORS

**Um S** conceived and designed the study, managed the data, conducted the statistical and geospatial analyses, interpreted the findings, and drafted the manuscript. **Chhea C** provided scientific supervision, contributed to the interpretation of the findings, and critically revised the manuscript. **Phy M** contributed to the study design, methodological guidance, interpretation of the findings, and critical revision of the manuscript. **Haruyama R** provided methodological guidance on geospatial analysis, contributed to the interpretation of the spatial findings, and critically revised the manuscript. All authors reviewed and approved the final version of the manuscript and agreed to be accountable for their respective contributions. **Um S** is the guarantor of the study and accepts responsibility for the overall content of the manuscript.

## PATIENT AND PUBLIC INVOLVEMENT

This manuscript reports a secondary analysis of an existing national survey and involved no new recruitment or interventions.

## DATA AVAILABILITY STATEMENT

The same de-identified dataset created by the Ministry of Health Cambodia has been sent to the WHO headquarters in September 2024, to be publicly available on the WHO NCD Microdata Repository (https://extranet.who.int/ncdsmicrodata/index.php/catalog/STEPS). As of September 2026, the dataset has yet to be shown in the repository; however, it can be shared by the Ministry of Health Cambodia on reasonable request.

---

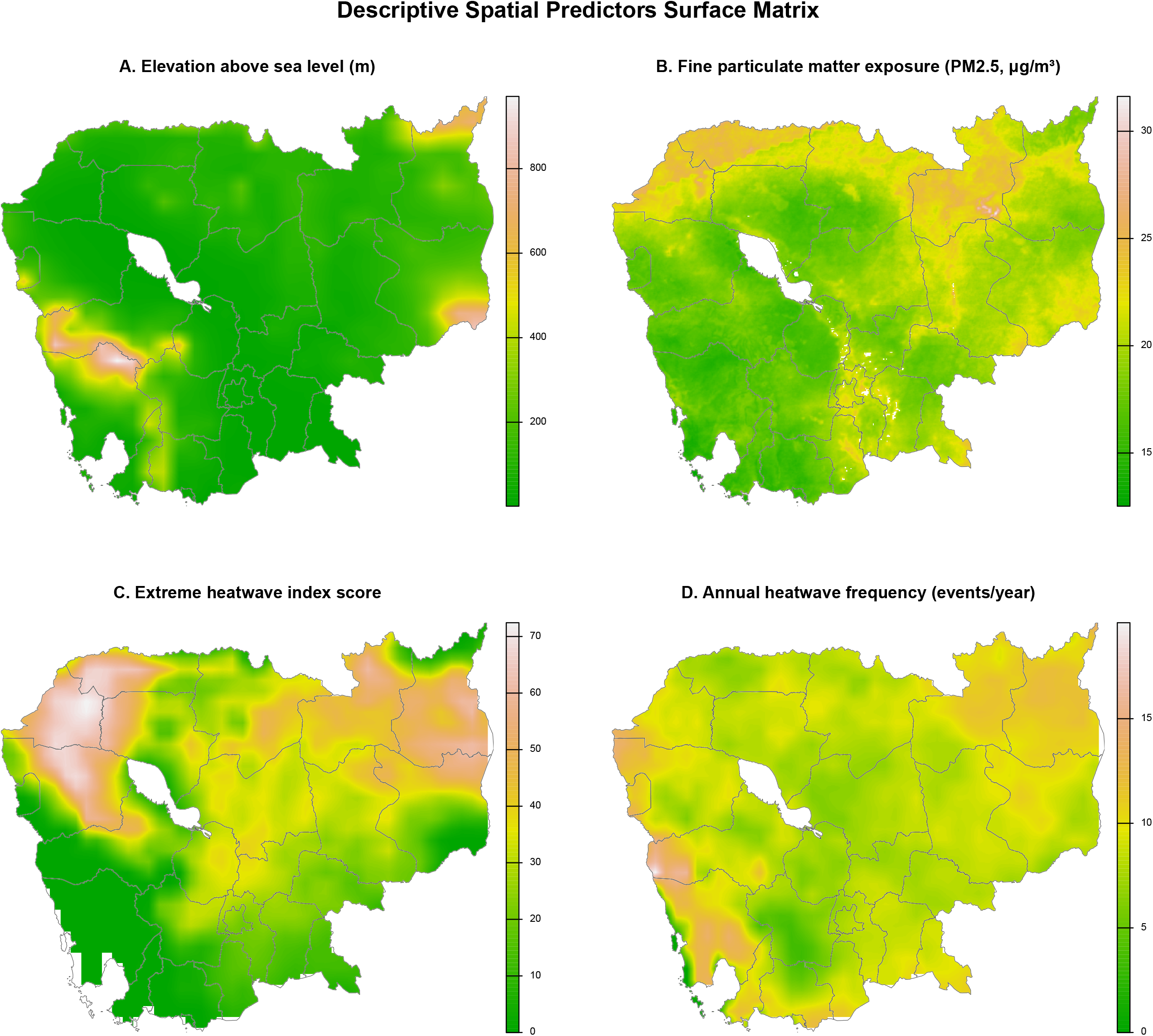

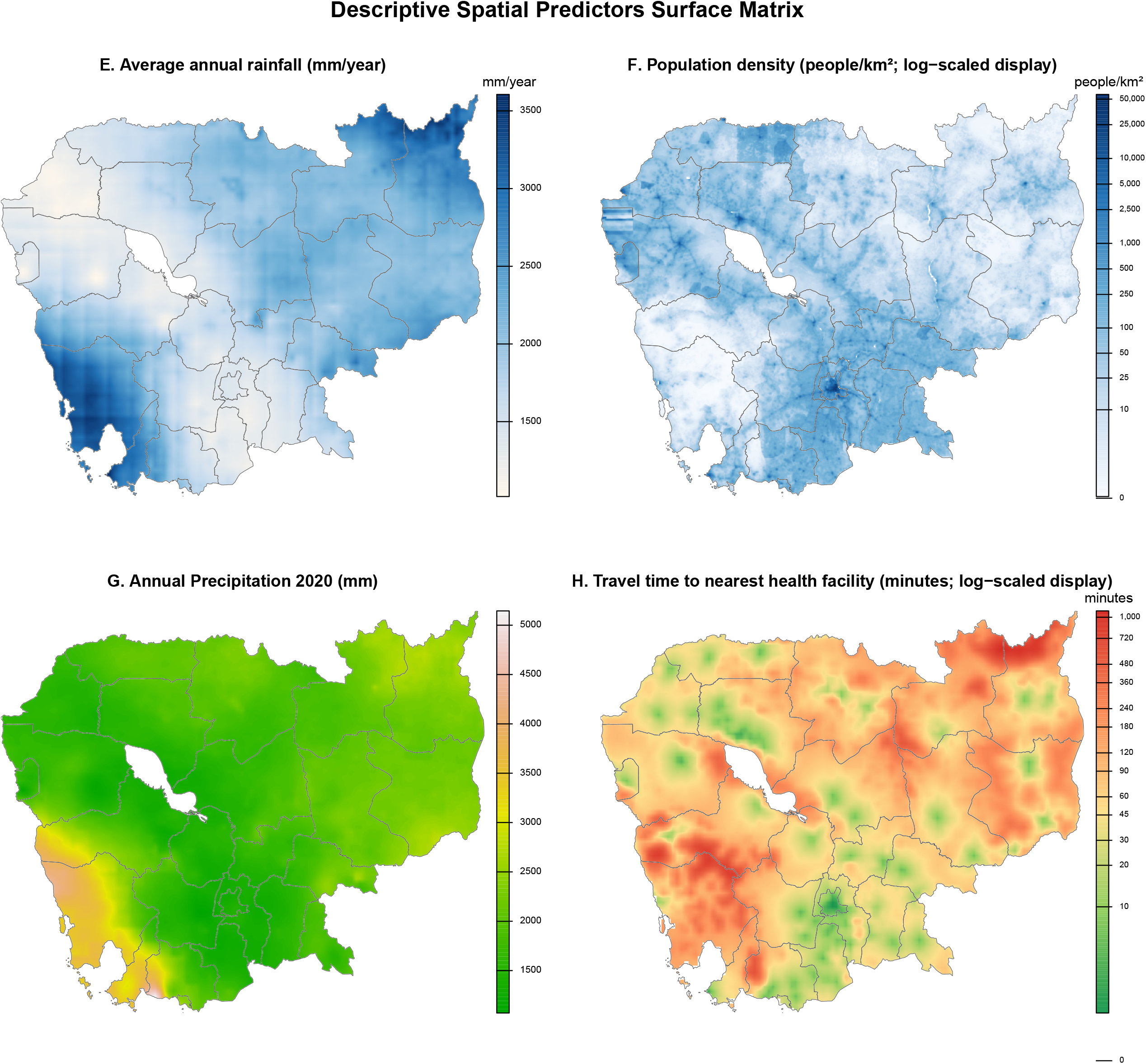

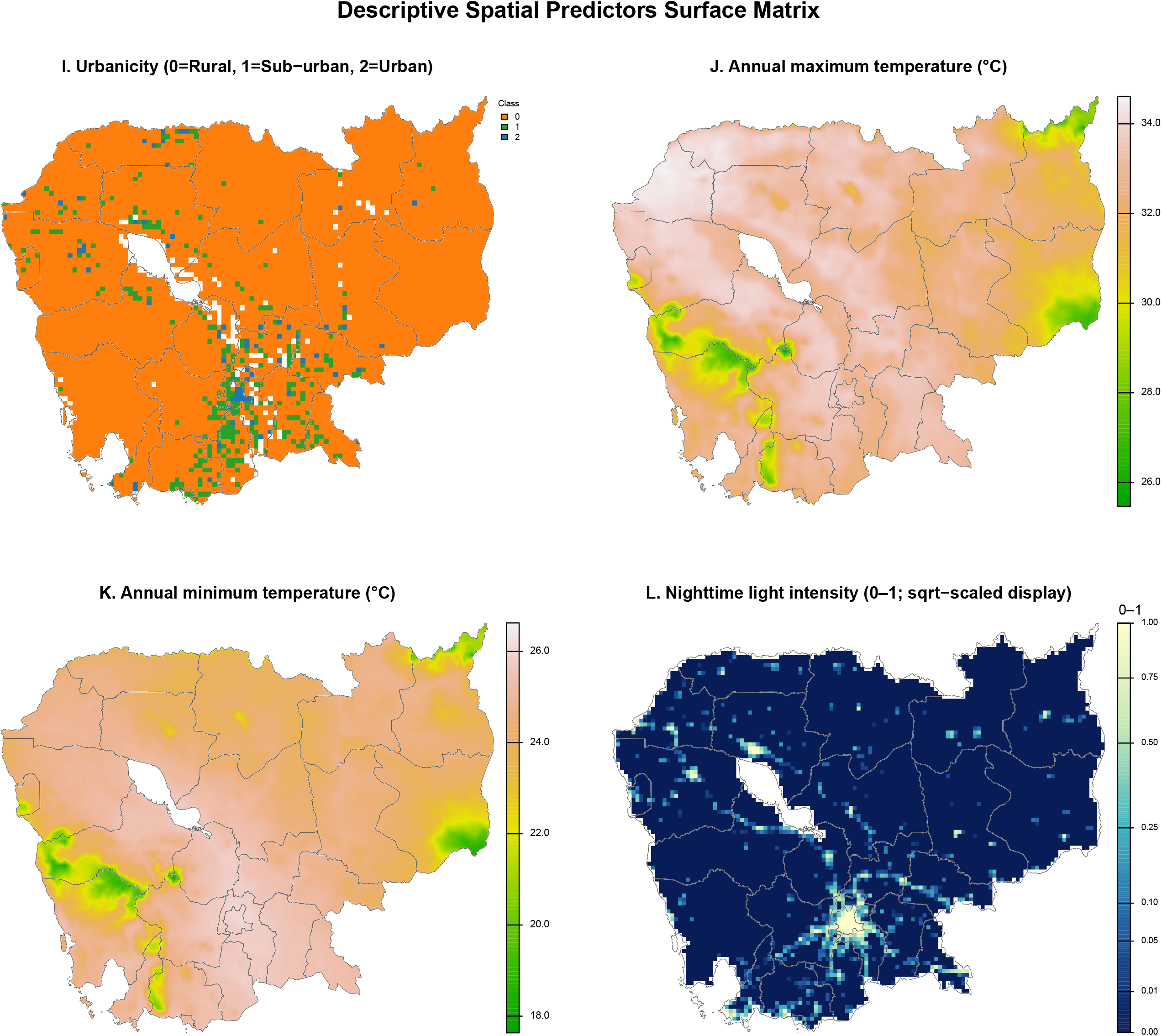

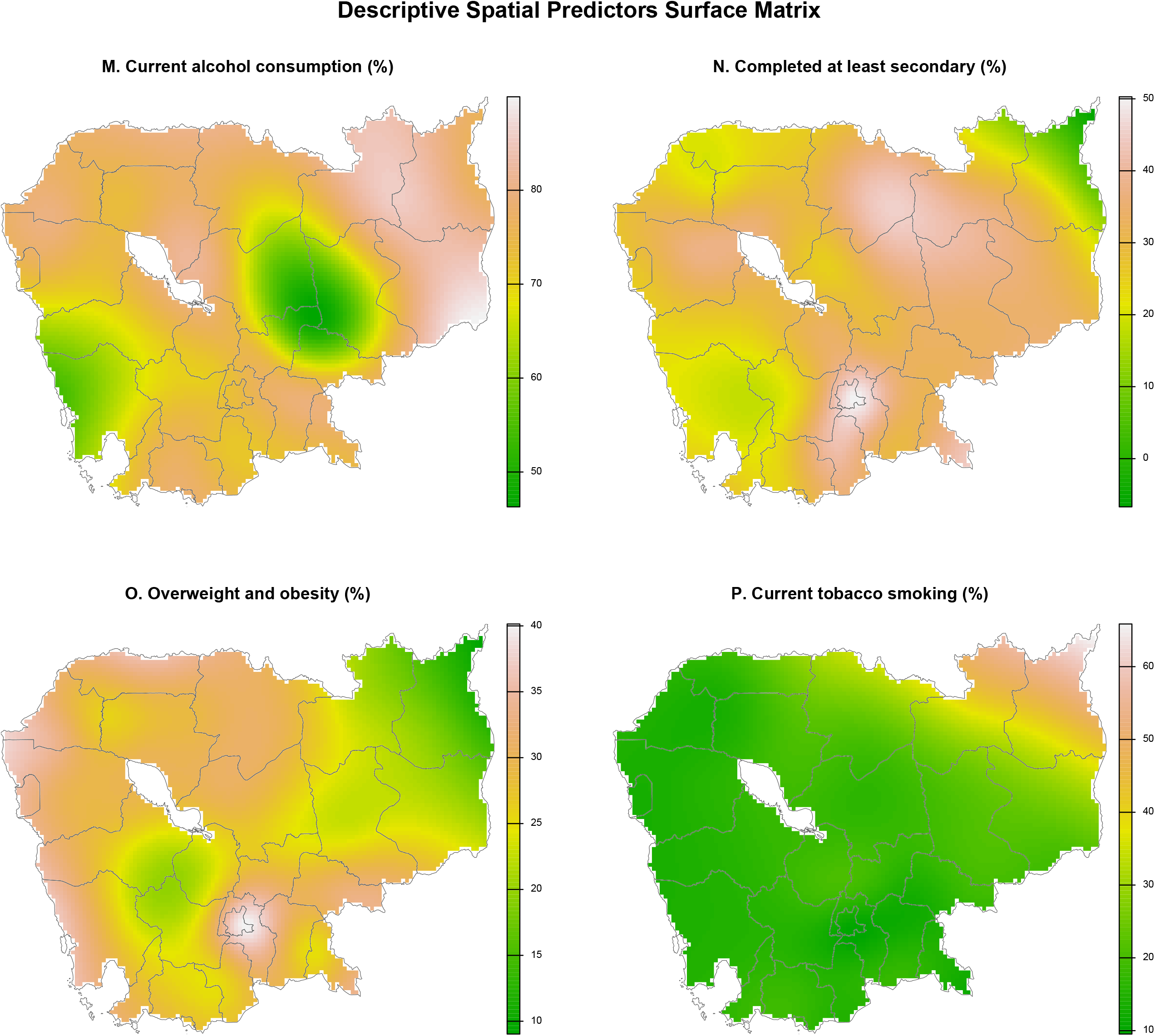

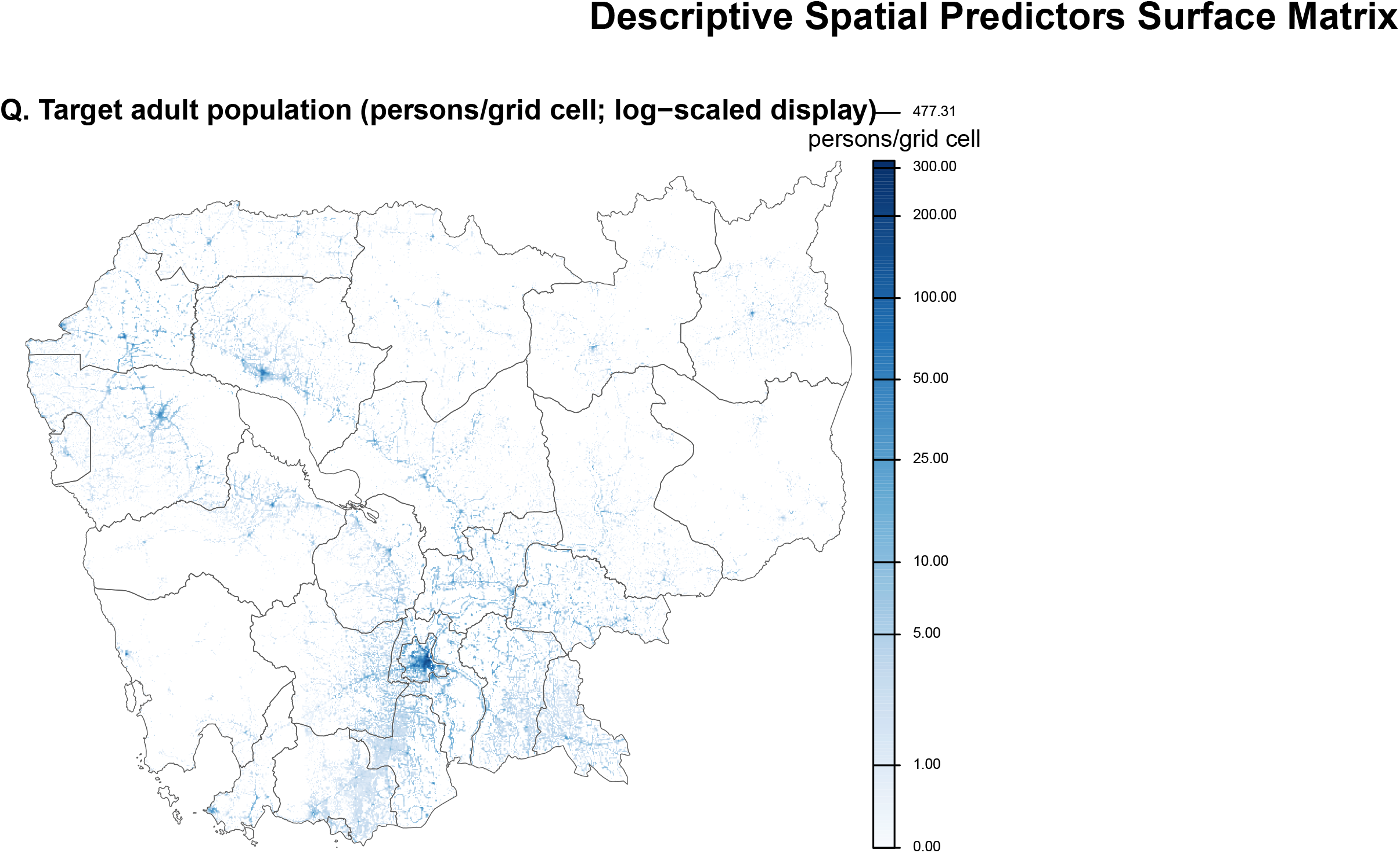

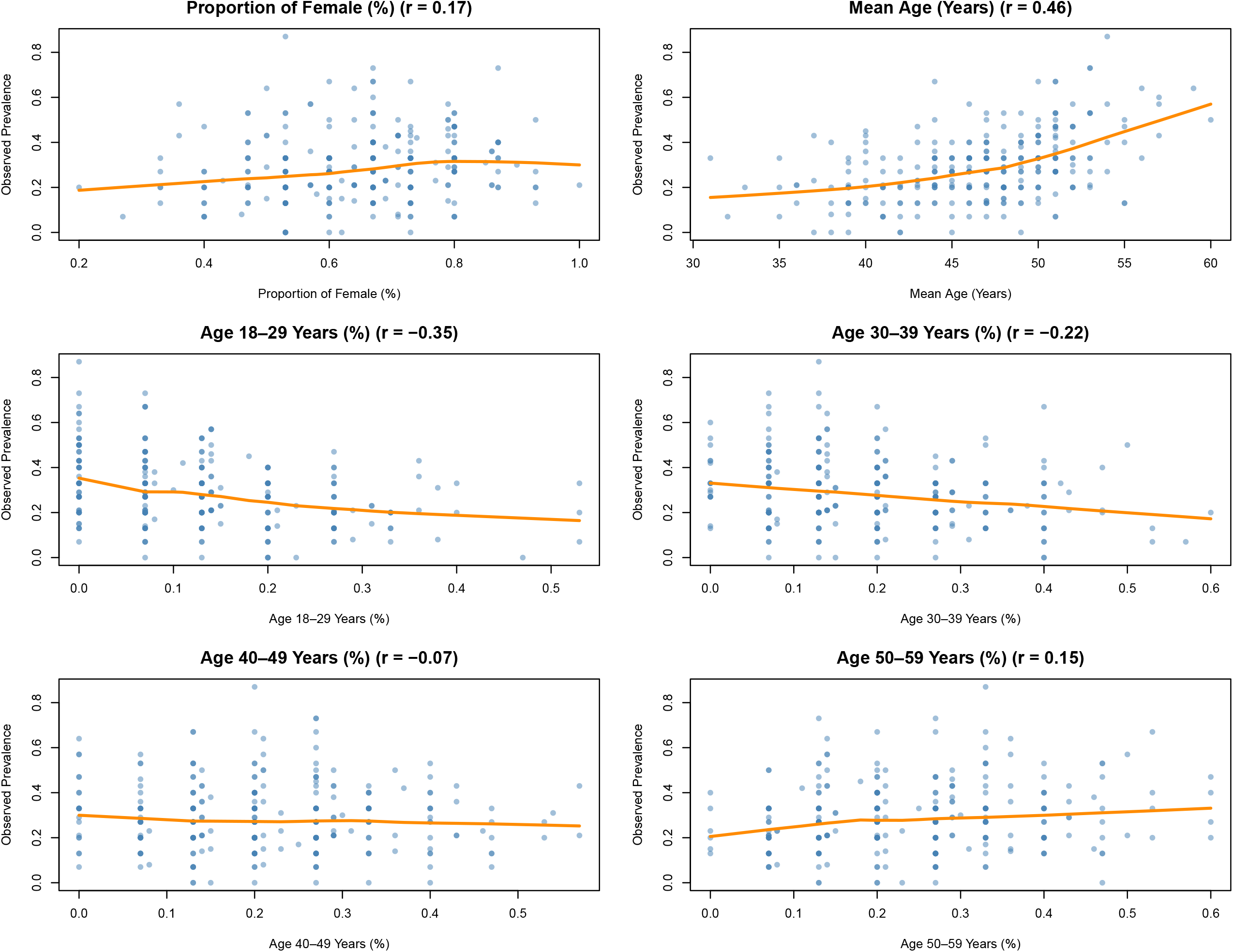

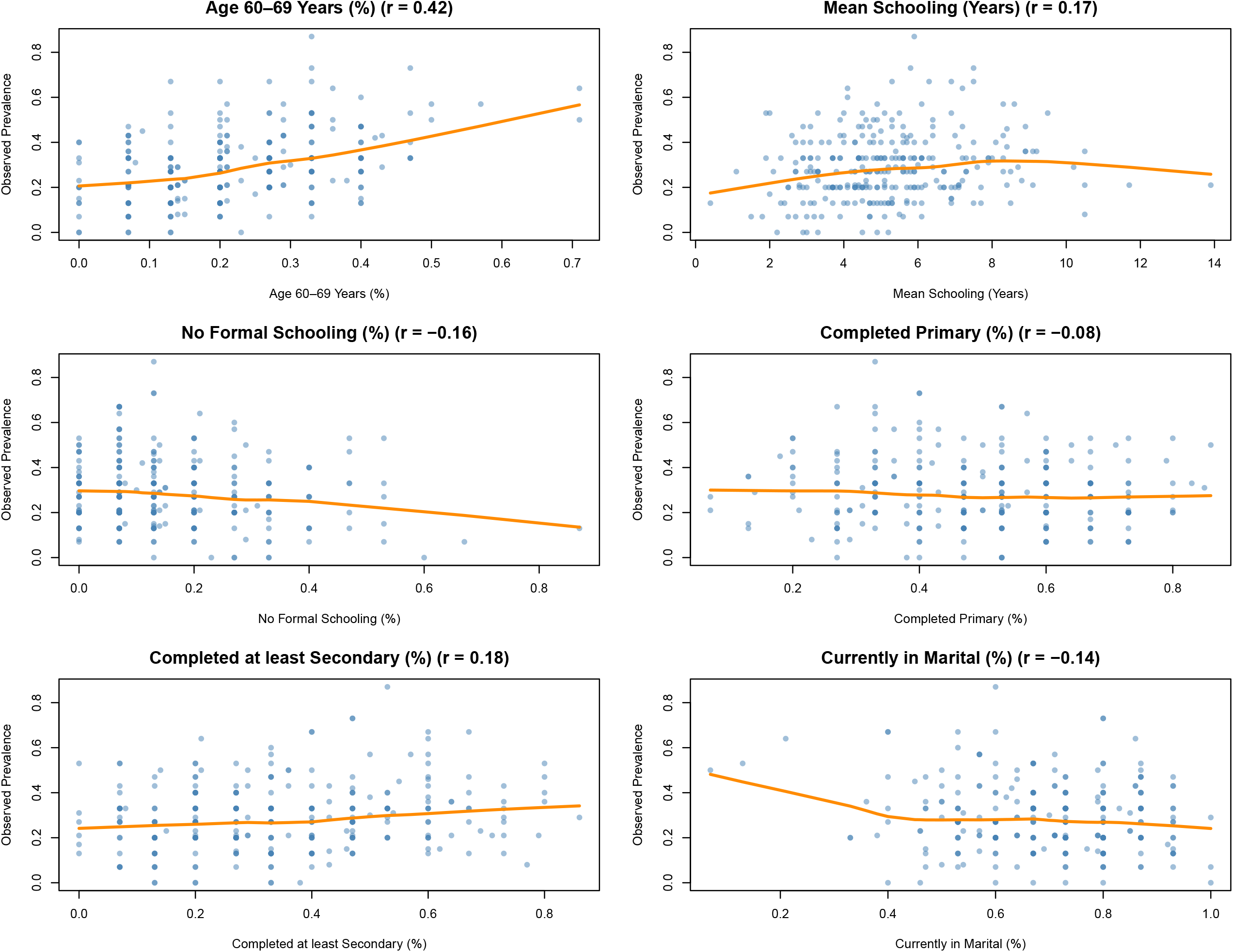

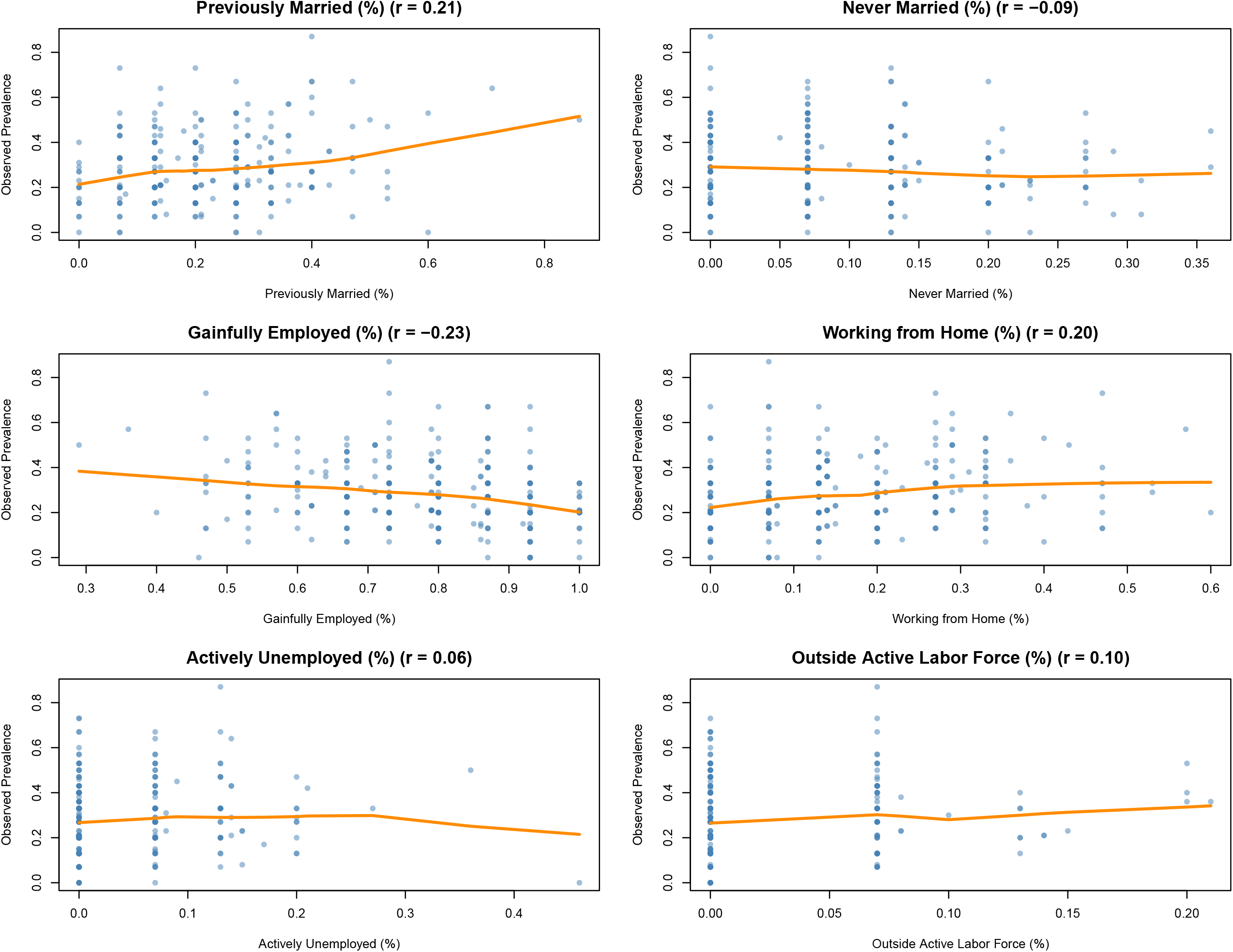

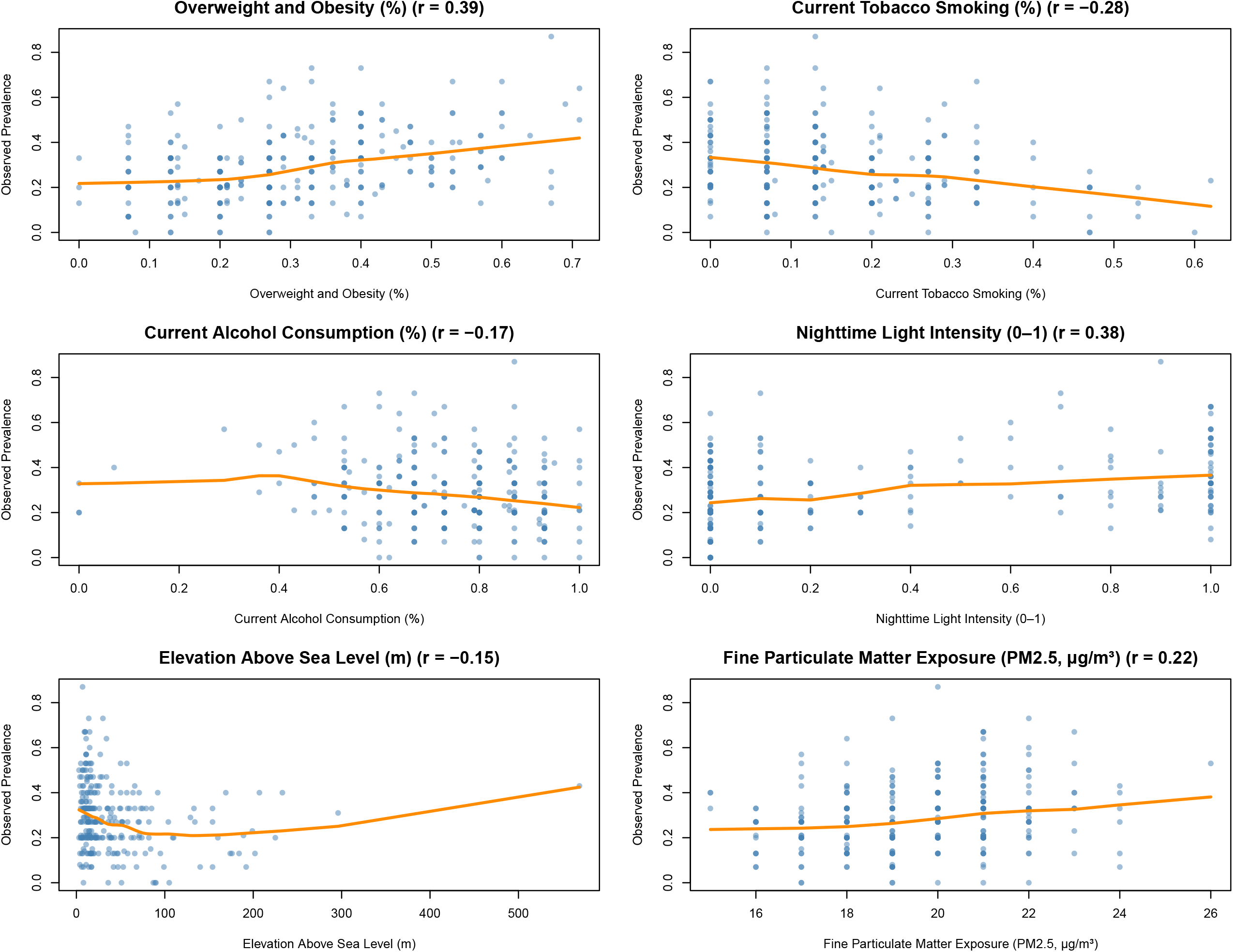

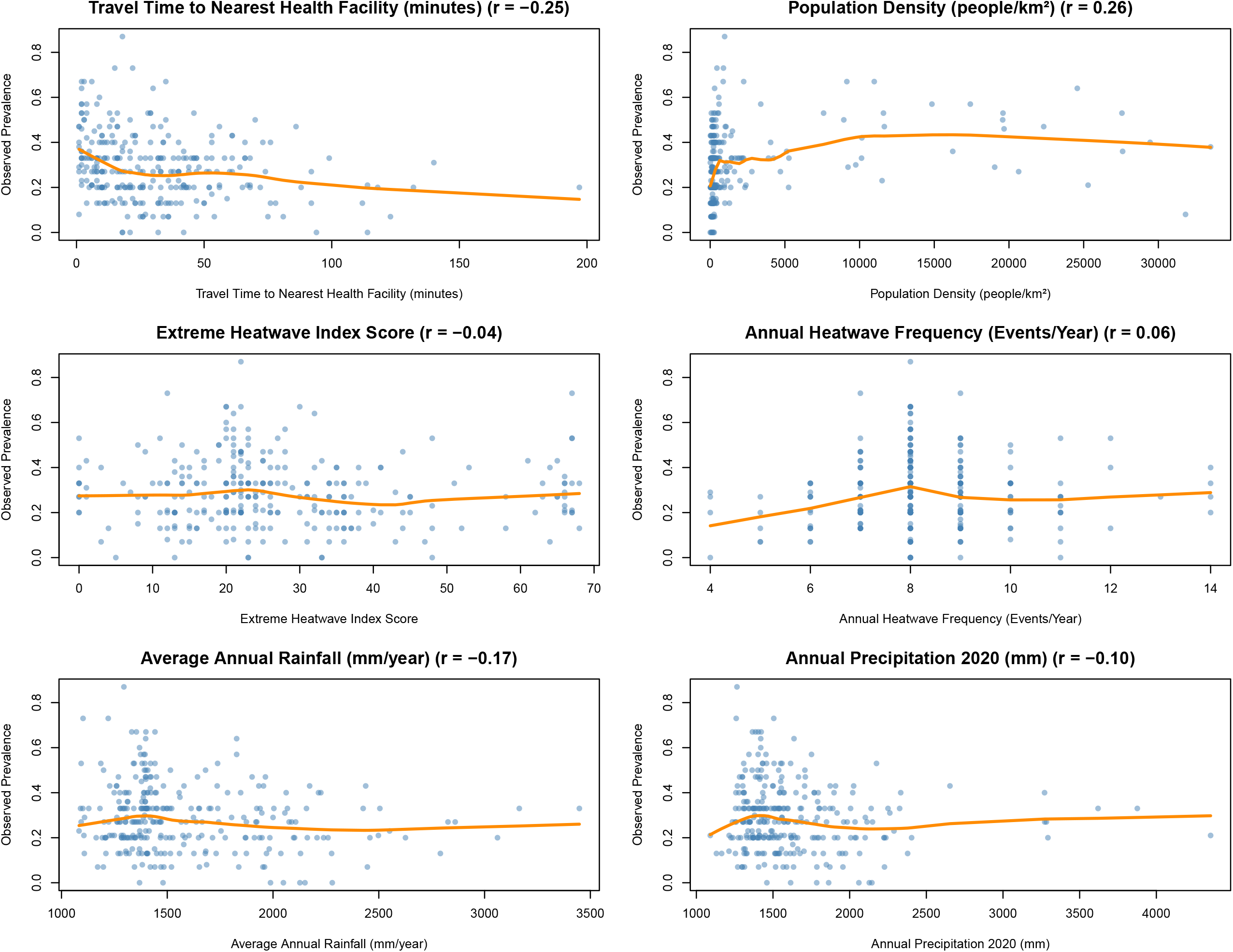

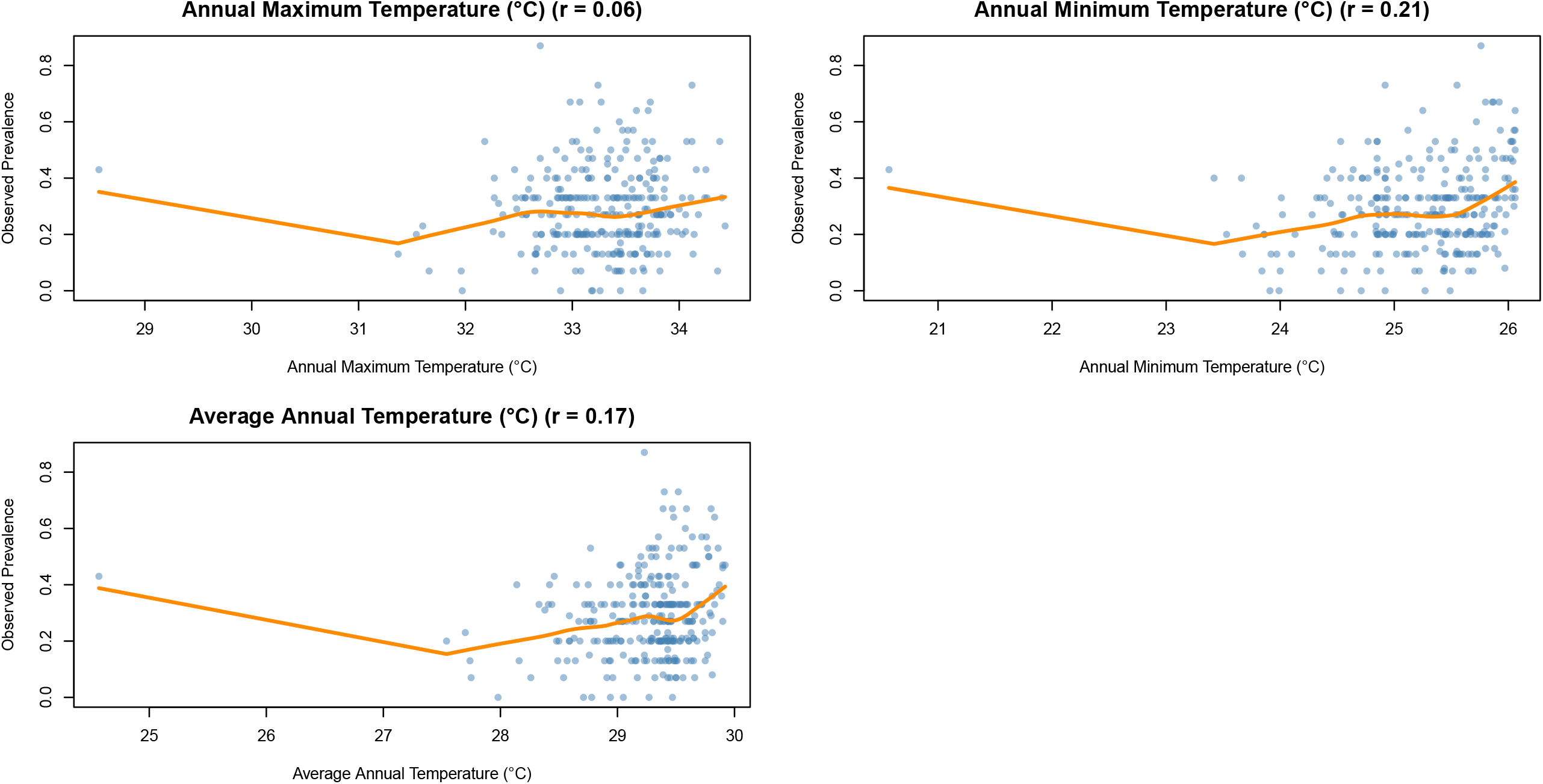

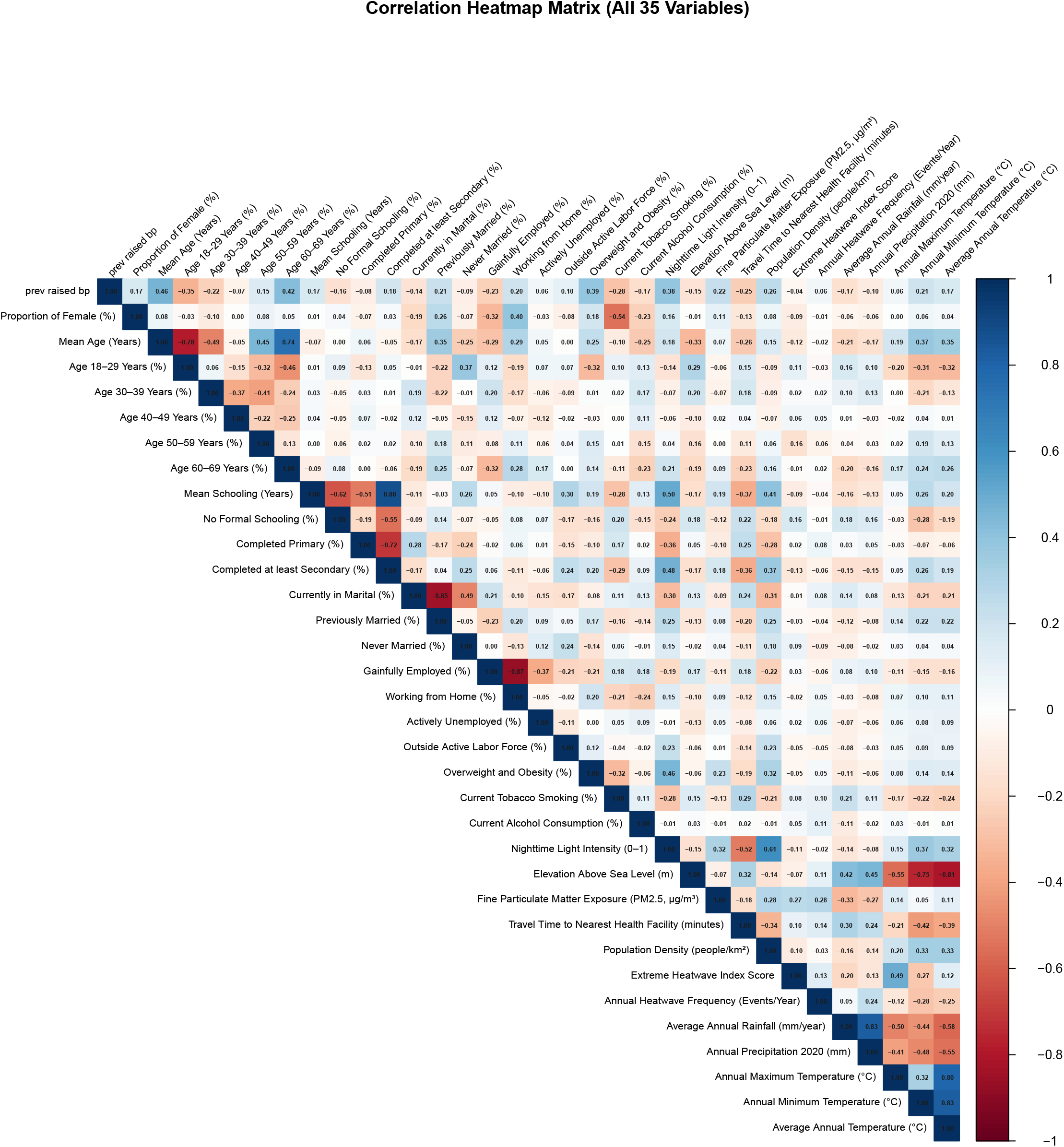

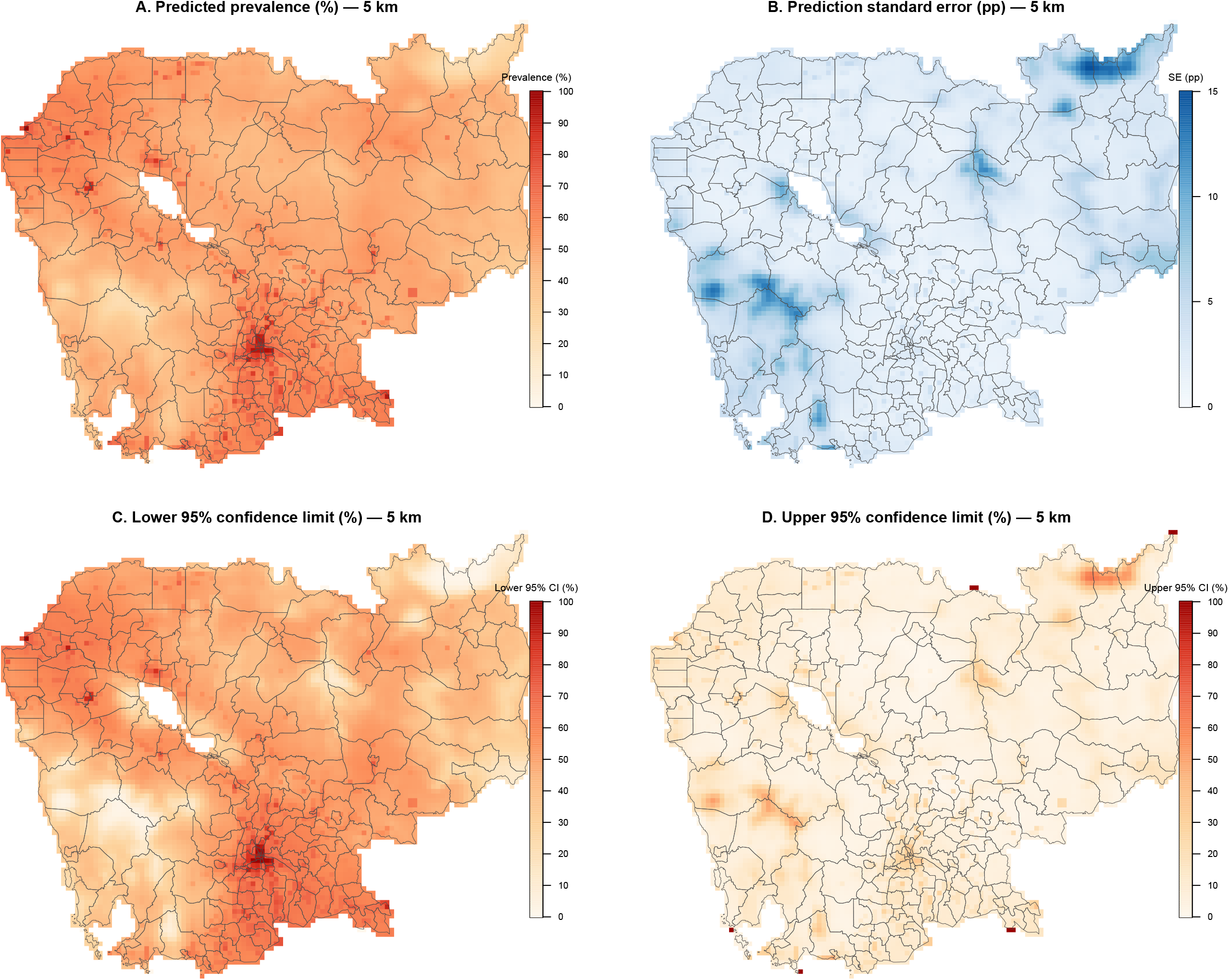

