## supplement for "Spatial variation and correlates of raised blood pressure in Cambodia: a Gaussian-process geostatistical analysis of the 2023 STEPwise survey"

**Online Supplementary Figure legends**

**Supplementary Figure S1.** Spatial distribution of candidate contextual covariates considered in the geospatial analysis of raised blood pressure in Cambodia, 2023. Maps show the spatial distribution of candidate environmental, climatic, built-environment and population covariates considered during model development: (**A**) elevation above sea level; (**B**) fine particulate matter (PM2.5); (**C**) extreme heatwave index; **(D**) annual heatwave frequency; (**E**) average annual rainfall; (**F**) population density; (**G**) annual precipitation; (**H**) travel time to the nearest health facility; (**I**) urbanicity; (**J**) annual maximum temperature; (**K**) annual minimum temperature; (**L**) nighttime-light intensity; (**M**) current alcohol consumption; (**N**) completion of at least secondary education; (**O**) overweight/obesity; (**P**) current tobacco smoking; and (**Q**) target adult population. Log or square-root transformations shown for selected highly skewed variables were used for visualisation only and did not alter the covariate values used in statistical modelling.

**Supplementary Figure S2.** Univariable relationships between observed PSU-level raised blood pressure prevalence and candidate covariates, Cambodia STEPS Survey 2023. Scatterplots show crude relationships between observed raised blood pressure prevalence at the primary sampling unit (PSU) level and candidate demographic, socioeconomic, behavioural, metabolic, built-environment and environmental covariates. Curves show locally weighted smooths, and r denotes the Pearson correlation coefficient. These analyses were used for exploratory assessment of direction, strength and potential non-linearity before multivariable modelling and should not be interpreted as adjusted or causal associations.

**Supplementary Figure S3.** Correlation matrix of observed PSU-level raised blood pressure prevalence and candidate covariates, Cambodia STEPS Survey 2023. Heatmap shows pairwise Pearson correlation coefficients between observed primary sampling unit (PSU)-level raised blood pressure prevalence and candidate demographic, socioeconomic, behavioural, metabolic, built-environment, climatic and environmental covariates. The matrix was used as an exploratory multicollinearity diagnostic to identify strongly correlated or potentially redundant representations before multivariable spatial modelling. Correlations represent crude pairwise relationships and should not be interpreted as independent or causal associations.

**Supplementary Figure S4**. Spatial distribution of model-predicted raised blood pressure prevalence and associated prediction uncertainty at 5-km resolution, with district administrative boundaries, Cambodia STEPS Survey 2023. (**A**) Predicted raised blood pressure prevalence (%); (**B**) prediction standard error (percentage points); (**C**) lower 95% confidence limit (%); and **(D**) upper 95% confidence limit (%). Predictions were generated using a binomial generalised additive model with a two-dimensional Gaussian-process spatial smooth. District boundaries are displayed for geographic reference; the maps represent 5-km grid-level predictions rather than independently estimated district-level prevalence.
