## supplement for "Spatial variation and correlates of raised blood pressure in Cambodia: a Gaussian-process geostatistical analysis of the 2023 STEPwise survey"

### Supplementary Table 1

Multicollinearity profiles across all 35 initial candidate predictors. VIF/GVIF diagnostics were used to identify redundant predictor representations before multivariable modelling; the diagnostics were not treated as an automatic causal or spatial selection rule.

| **Candidate Covariate** | **Generalized VIF Score** | **Model Inclusion Threshold Status** | **Pipeline Threshold Enforcement Action** |
| --- | --- | --- | --- |
| Proportion of Female (%) | 1.90 | Safe Baseline (< 5.0) | Retained in adjusted model |
| Mean Age (Years) | 37.83 | Highly Collinear (>= 5.0) | Omitted Pre-estimation |
| Age 18–29 Years (%) | 31.13 | Highly Collinear (>= 5.0) | Omitted Pre-estimation |
| Age 30–39 Years (%) | 22.51 | Highly Collinear (>= 5.0) | Omitted Pre-estimation |
| Age 40–49 Years (%) | 12.19 | Highly Collinear (>= 5.0) | Omitted Pre-estimation |
| Age 50–59 Years (%) | 4.50 | Safe Baseline (< 5.0) | Omitted Pre-estimation |
| Age 60–69 Years (%) | 1.00 | Safe Baseline (< 5.0) | Retained in adjusted model |
| Mean Schooling (Years) | 6.53 | Highly Collinear (>= 5.0) | Retained in adjusted model |
| No Formal Schooling (%) | 1.94 | Safe Baseline (< 5.0) | Retained in adjusted model |
| Completed Primary (%) | 1.00 | Safe Baseline (< 5.0) | Omitted Pre-estimation |
| Completed at least Secondary (%) | 5.10 | Highly Collinear (>= 5.0) | Retained in adjusted model |
| Currently in Marital (%) | 5.40 | Highly Collinear (>= 5.0) | Omitted Pre-estimation |
| Previously Married (%) | 5.39 | Highly Collinear (>= 5.0) | Omitted Pre-estimation |
| Never Married (%) | 1.00 | Safe Baseline (< 5.0) | Omitted Pre-estimation |
| Gainfully Employed (%) | 15.28 | Highly Collinear (>= 5.0) | Retained in adjusted model |
| Working from Home (%) | 13.67 | Highly Collinear (>= 5.0) | Omitted Pre-estimation |
| Actively Unemployed (%) | 3.76 | Safe Baseline (< 5.0) | Omitted Pre-estimation |
| Outside Active Labor Force (%) | 1.00 | Safe Baseline (< 5.0) | Retained in adjusted model |
| Overweight and Obesity (%) | 1.64 | Safe Baseline (< 5.0) | Retained in adjusted model |
| Current Tobacco Smoking (%) | 2.02 | Safe Baseline (< 5.0) | Retained in adjusted model |
| Current Alcohol Consumption (%) | 1.32 | Safe Baseline (< 5.0) | Retained in adjusted model |
| Nighttime Light Intensity (0–1) | 3.80 | Safe Baseline (< 5.0) | Retained in adjusted model |
| Elevation Above Sea Level (m) | 4.36 | Safe Baseline (< 5.0) | Retained in adjusted model |
| Fine Particulate Matter Exposure (PM2.5, ug/m3) | 1.64 | Safe Baseline (< 5.0) | Retained in adjusted model |
| Travel Time to Nearest Health Facility (per 10 min) | 1.76 | Safe Baseline (< 5.0) | Retained in adjusted model |
| Population Density (people/km²) | 3.11 | Safe Baseline (< 5.0) | Retained in adjusted model |
| Extreme Heatwave Index Score | 2.41 | Safe Baseline (< 5.0) | Omitted Pre-estimation |
| Annual Heatwave Frequency (Events/Year) | 1.72 | Safe Baseline (< 5.0) | Retained in adjusted model |
| Average Annual Rainfall (per 100 mm) | 5.66 | Highly Collinear (>= 5.0) | Retained in adjusted model |
| Annual Precipitation 2020 (mm) | 5.59 | Highly Collinear (>= 5.0) | Omitted Pre-estimation |
| Annual Maximum Temperature (°C) | 8174.36 | Highly Collinear (>= 5.0) | Omitted Pre-estimation |
| Annual Minimum Temperature (°C) | 9629.17 | Highly Collinear (>= 5.0) | Omitted Pre-estimation |
| Average Annual Temperature (°C) | 23529.89 | Highly Collinear (>= 5.0) | Omitted Pre-estimation |

### Supplementary Table 2

Model-fit diagnostics comparing the non-spatial explanatory GLM and spatial explanatory GAM using the same fixed-effect structure.

| **Statistical Fit Paradigm** | **Non-Spatial Model (Baseline GLM)** | **Model-Based Geostatistical (MBG) Framework** |
| --- | --- | --- |
| Akaike Information Criterion (AIC) | 1177.67 | 1151.45 |
| Bayesian Information Criterion (BIC) | 1203.29 | 1237.60 |
| Residual Deviance Metric | 362.20 | 302.89 |
| Deviance Explained (%) | 15.8% | 37.7% |

.
